# The impact of adaptive mosquito behavior and insecticide-treated nets on malaria prevalence

**DOI:** 10.1101/2020.05.18.20105916

**Authors:** Calistus N. Ngonghala, Josephine Wairimu, Jesse Adamski, Hardik Desai

**Affiliations:** Department of Mathematics, University of Florida, Gainesville, FL 32611, USA; Emerging Pathogens Institute, University of Florida, Gainesville, FL 32608, USA; Center for African Studies, University of Florida, Gainesville, FL 32611, USA; School of Mathematics, University of Nairobi, Kenya

**Keywords:** Insecticide-treated net coverage and efficacy, Mosquito resistance, Malaria prevalence, Malaria control, Mosquito behavior

## Abstract

Malaria prevalence in sub-Saharan Africa remains high. Kenya for example, records about 3.5 million new cases and 11 thousand deaths each year [1]. Most of these cases and deaths are among children under five. The main control method in malaria endemic regions has been through the use of pyrethroid-treated bed nets. Although this approach has been fairly successful, the gains are threatened by mosquito-resistance to pyrethroids, physical and chemical degradation of ITNs that reduce their efficacy, inconsistent and improper use by humans, etc. We present a model to investigate the effects of insecticide-treated bed-net use and mosquito-resistance and adaptation to pyrethroids used to treat bed nets on malaria prevalence and control in malaria endemic regions. The model captures the development and loss of resistance to insecticides, the effects of bed-net use on malaria control in a setting where proper and consistent use is not guaranteed, as well as differentiated biting of human hosts by resistant and sensitive mosquitoes. Important thresholds, including the basic reproduction number *R*_0_, and two parameter groupings that are important for disease control and for establishing the existence of endemic equilibria to the model are calculated. Furthermore, a global sensitivity analysis is carried out to identify important parameters such as insecticide treated bed-net coverage, insecticide treated bed-net efficacy, the maximum biting rate of resistant mosquitoes, etc., that drive the system and that can be targeted for disease control. Threshold levels of bed-net coverage and bed-net efficacy required for containing the disease are identified and shown to depend on the type of insecticide-resistance. For example, when mosquito-resistance to insecticides is not permanent and is acquired only through recruitment and the efficacy of insecticide-treated nets is 90%, about 70% net coverage is required to contain malaria. However, for the same insecticide-treated net efficacy, i.e., 90%, approximately 93% net coverage is required to contain the disease when resistance to insecticides is permanent and is acquired through recruitment and mutation in mosquitoes. The model exhibits a backward bifurcation, which implies that simply reducing *R*_0_ slightly below unity might not be enough to contain the disease. We conclude that appropriate measures to reduce or eliminate mosquito-resistance to insecticides, ensure that more people in endemic areas own and use insecticide-treated nets properly, and that the efficacy of these nets remain high most of the times, as well as educating populations in malaria endemic areas on how to keep mosquito densities low and minimize mosquito bites are important for containing malaria.

## 1. Introduction

Malaria is a vector-borne disease caused by *Plasmodium* parasites and spread by infectious female mosquitoes as they seek blood required for development of their eggs. There are four major types of human malaria parasites: *Plasmodium falciparum, Plasmodium vivax, Plasmodium malariae*, and *Plasmodium ovale. Plasmodium falciparum* and *Plasmodium vivax* are the most common forms with *Plasmodium falciparum* the most deadly type, especially in sub-Saharan Africa, where it causes more than 400,000 deaths each year [2]. More than 90% of malaria-imposed deaths occur in Africa and mostly amongst children under the age of five. The disease costs economies in endemic areas thousands of dollars in treatment, control, management and elimination efforts each year. For example, it is estimated that US $12 billion is spent annually to fight malaria [3]. In Sub-Saharan Africa, there are two main malaria transmitting vector species–*Anopheles gambiae* and it Anopheles arebiansis. *Anopheles gambiae* is the world’s most effective vector of human malaria because of its susceptibility to *Plasmodium* parasites, short development time, preference for human-blood, and preference for indoor-feeding and resting [4].

The fight against endemic malaria in Sub-Saharan Africa has been stepped up within the past 15 years. Most efforts have been directed towards vector control measures, which include provision and use of Insecticide Treated Nets (ITNs) (i.e., Long Lasting Insecticide Nets (LLINs) and treated regular nets) and Indoor Residual Spraying (IRS) with insecticides, which target *Anopheles* malaria vectors. Insecticide treated nets and indoor residual spraying use synthetic chemicals called pyrethroid insecticides that kill and repel mosquitoes. These two vector intervention strategies account for 25% of the total world pyrethroids market [5]. They are widely used in Kenya, especially in regions with increased malaria prevalence and vector resistance, e.g., the Western and Coastal regions of Kenya, where malaria is endemic and is transmitted year-round with increased transmission. Malaria prevalence in these areas remains high despite the control efforts made over the years [6]. The fight against the disease in Kenya spans several decades with enhanced strategies and concerted donor country efforts yielding successful results, although only to a limited extent. Different regions report success of different control strategies ranging from awareness campaigns on the role of mosquitoes in malaria transmission, reduction of mosquito breeding sites near homes, personal and family protection through proper use of ITNs and mosquito repellents, through indoor residual spraying [4, 7]. One common observation has been that sustained vector control is a key intervention measure for any control progress to translate into the ambitious goal of malaria elimination [8–10].

Since malaria affects mostly the rural poor populations, ITNs have been very useful in reducing morbidity and mortality in these populations due to their low cost and ease in implementation [11]. Distribution of free ITNs to the most vulnerable group of humans, i.e., pregnant women and children below five has seen malaria incidence decline in Africa [1, 12–15]. In particular, ITNs have been responsible for averting approximately 68% of malaria-related deaths in Africa [16]. Insecticide-treated nets are about 90% effective when new, however, due to several factors including natural wear and misuse, the efficacy can drop to less than 70% over the lifespan (three years). Untreated bed-nets that are in good condition or ITNs that have lost their efficacy provide about 50% protection to humans [17]. Insecticide-treated net coverage in Kenya, for example, has increased drastically from 7% in 2004 through 67% in 2006 [18], to over 80% in 2015 [19]. To realize the expected reduction in malaria transmission through the use of ITNs, ITN efficacy and coverage for at risk populations must be high enough [11]. Unfortunately, low coverage and low efficacy coupled with differentiated adherence to the use of ITNs has a negative impact on malaria control [15]. The aim of universal ITN coverage is to attain a 1:1.6 ratio in order to reduce malaria prevalence to an acceptable level. But, achieving this goal is hampered by several challenges including ITN ownership, regular wear, misuse, inconvenience (e.g., people tend to sleep out of ITNs when it is hot), human behavior, perception, and literacy level, etc. [19–23]. These confounding factors make ITN coverage and efficacy variable in endemic areas. Understanding the impact of these limitations can help us assess the effectiveness of ITNs, devise optimal control strategies using these ITNs, and guide public health policy.

Another challenge to the gains from vector control measures against malaria such as ITNs is resistance exhibited by mosquitoes to pyrethroid insecticides used in ITNs [24–26]. Insecticide-resistance is defined as the increased ability of insects to withstand or overcome the toxic, killing, or repellent effects of insecticides through natural selection and mutation. Thus, resistance is measured by the effectiveness of insecticides in killing mosquitoes, as well as the ability of some vectors to tolerate the toxic effects. For example, when mosquitoes are exposed to insecticides, the resistance is low if the mosquitoes have a 0-40% survival probability, medium if they have a 40-80% survival probability, and high if the mosquito survival probability is at least 80% [27]. Mosquitoes respond to insecticide exposure behaviorally, numerically, or evolutionarily [28]. Behavioral response involves mosquitoes backing off from toxic sprays or ITNs without biting to return for a blood meal only after active ingredients in the insecticides have subsided [29–31]. Numeric response has resulted in a decline in the population of mosquitoes, as well as a shortened lifespan of mosquitoes [32–34]. Evolutionary changes occur when there is reduced sensitivity to insecticides in ITNs. In this case, there is target site blocking and increased frequency of metabolism [35–37].

Many types of mosquito resistance to insecticides have been identified. These include behavioral, metabolic, and cuticular resistance. Behavioral resistance occurs when mosquitoes adapt to human protective behavior. For example, mosquitoes might bite earlier before humans sleep under ITNs and rest outside sprayed human homes to avoid the toxicity of pyrethroids. Metabolic resistance occurs when mosquitoes undergo several mutations (causing changes to the chemical target site), which enable them to detoxify the chemical or withstand prolonged exposure to insecticides without being killed [38–41]. Cuticular resistance is a physical characteristic of mosquitoes, where their cuticles are thickened and the composition of the cuticle is altered in order to absorb less insecticide [42, 43]. This can result in increased resistance to insecticides [44]. Although behavioral resistance can be acquired and lost depending on survival from exposure, metabolic and cuticular resistance are permanent, i.e. a metabolic or cuticle resistant mosquito maintains its resistance until it dies. Mosquito-resistance to insecticides is widely distributed [45] and its intensity is increasing, thus reducing the effectiveness of ITNs and other pyrethroid-related malaria control measures [28, 45]. To make matters worst, multiple resistance to insecticides have been detected in most of the countries in which malaria is endemic [46]. For example, when multiple pyrethroid-related interventions, e.g., ITN, IRS, and mosquito-repellents are used, about 3-5% of the infectious mosquitoes develop resistance [47]. The emerging increase in resistance levels coupled with normal wear, misuse of ITNs, and human behavior is a major threat to the efficacy of protection from ITNs [26]. This calls for extensive studies to understand and monitor the effects of long-term implementation of vector control measures, the emergence of insecticide-resistance and the effects of resistance on the efficacy of these control measures [28, 29, 48]. This information is important for identifying and implementing better and more effective control measures [48].

One approach that has been useful for gaining insights into the complex processes that surround the persistence of malaria is mathematical modeling (see, for example, [49–63]). Some mathematical models have focused on understanding the role of ITNs, e.g., [60, 64–69] on malaria control, while others have focused on investigating the effects of mosquito-resistance to insecticides on malaria dynamics [70–74]. Here, we investigate the impact of ITNs and the extent to which insecticide resistance affect malaria control efforts in endemic areas. We approach this through a mathematical model that incorporates ITN-use and two types of mosquitoes–sensitive and resistant mosquitoes. To our knowledge, this is the first mathematical model framework for malaria dynamics that combines ITN-use and the impact of mosquito resistance on the efficacy of ITNs to understand malaria prevalence and control.

## 2. Model formulation

We develop the mathematical model in this section. Key features of the model include explicit incorporation of resistance to insecticides by mosquitoes and personal protection through the use of insecticide-treated bed nets. Schematics for the model system are presented in Fig. 1. We consider a Susceptible-Infectious-Partially immune-Susceptible (SIRS) framework for the human population. That is the total human population denoted by *N*_*h*_ is divided into three compartmental classes–susceptible (humans who do not have malaria) denoted by *S*_*h*_, infectious (humans who have malaria and can transmit it) denoted by *I*_*h*_, and partially immune (humans who have temporary immunity to malaria) denoted by *R*_*h*_. The susceptible human population is increased by natural births that occur at rate Λ_*h*_ (we are assuming that malaria is not vertically transmissible) and when partially immune humans lose immunity at rate *ρ*_*h*_. The population of this class is decreased through natural deaths that occur at per capita *µ*_*h*_ and through new infections from infectious mosquitoes modeled through the force of infection *λ*_*vh*_. The infectious human population increases through new infections and decreases when humans die naturally at per capita rate *µ*_*h*_, acquire partial-immunity at per capita rate *γ*_*h*_, or are killed by the disease at per capita rate *δ*_*h*_. The partially immune human population is increased by newly immune humans from the infectious class and reduced by natural mortalities and through loss of immunity at per capita rate, *ρ*_*h*_. Based on this description and the schematics presented in Fig. 1, the human population and disease dynamics within the human population are described by the system of equations:

**Figure 1:**
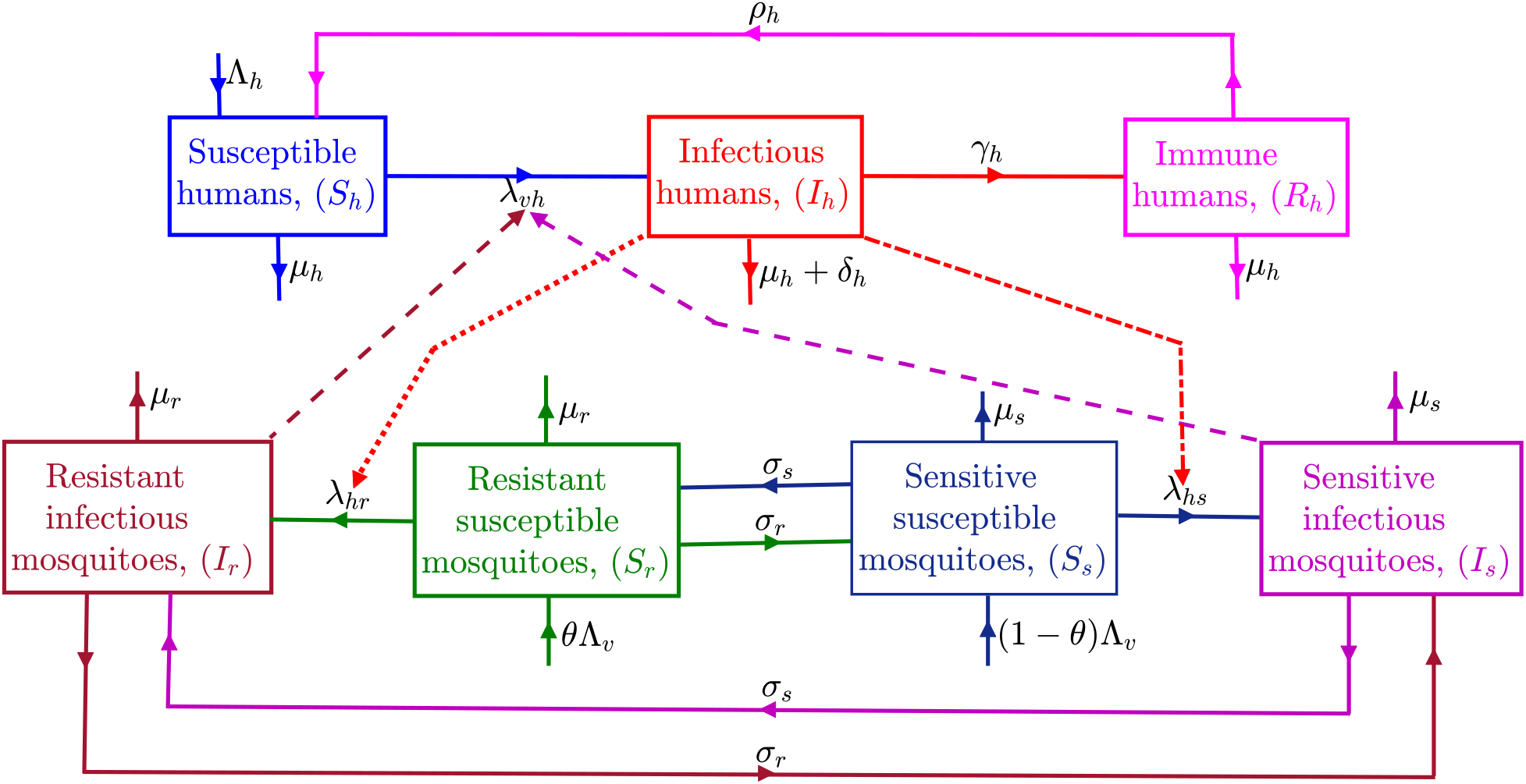
Model flow-chart showing the flow of humans and mosquitoes between different classes that represent the status of the disease (solid lines) and interactions between humans and mosquitoes (non solid lines). Interactions resulting in sensitive and resistant susceptible mosquito infections by infectious humans are denoted by dash-doted and dotted lines, respectively, and interactions resulting in susceptible human infections by sensitive and resistant infectious mosquitoes are denoted by dark red and light magenta dashed lines, respectively. The human population is broken down into susceptible *S*_*h*_, infectious *I*_*h*_, and partially immune *R*_*h*_, while the mosquito population comprises susceptible sensitive and resistant (*S*_*s*_ and *S*_*r*_, respectively,) and infectious sensitive and resistant classes (*I*_*s*_ and *I*_*r*_, respectively). Descriptions of the transition rates (parameters) are presented in the text and in Table 1, while the forces of infection *λ*_*vh*_, *λ*_*hs*_ and *λ*_*hr*_ are described in the text and presented in Eq. (2.6).

**Figure 2:**
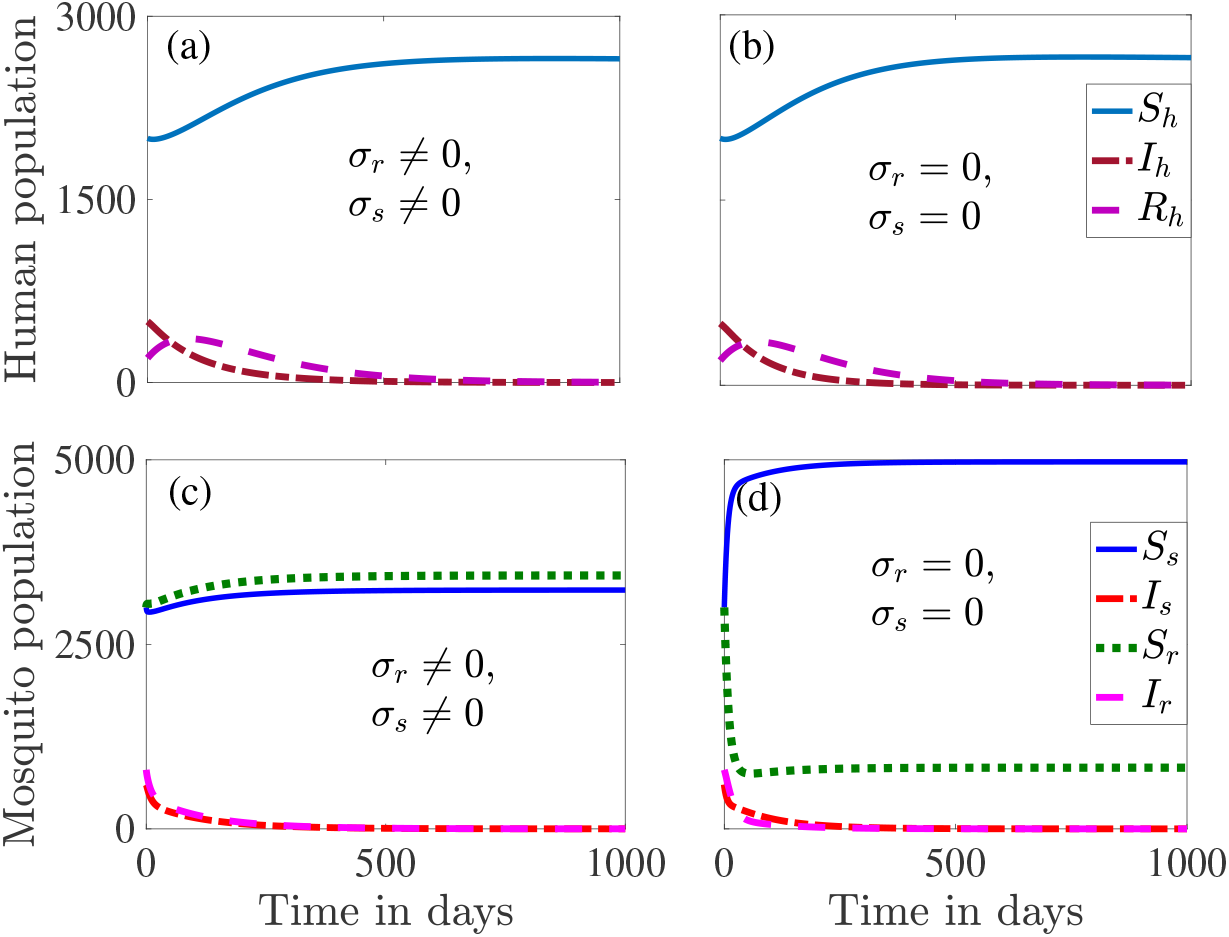
Numerical simulation results illustrating the existence of a disease free equilibrium to System (2) when *R*_0_ < 1. Graphs (a) and (c) show the dynamics for the case in which resistance is acquired at rate *σ*_*s*_ ≠ 0 and lost over time at rate *σ*_*r*_ ≠ 0. Graphs (b) and (d) illustrate the dynamics when resistance is acquired only through mosquito recruitment and is permanent. Parameter values used for the simulations are presented in Table 1.

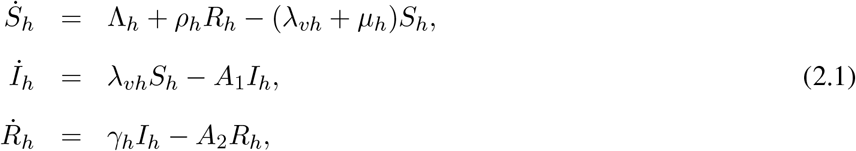

**Table 1:**
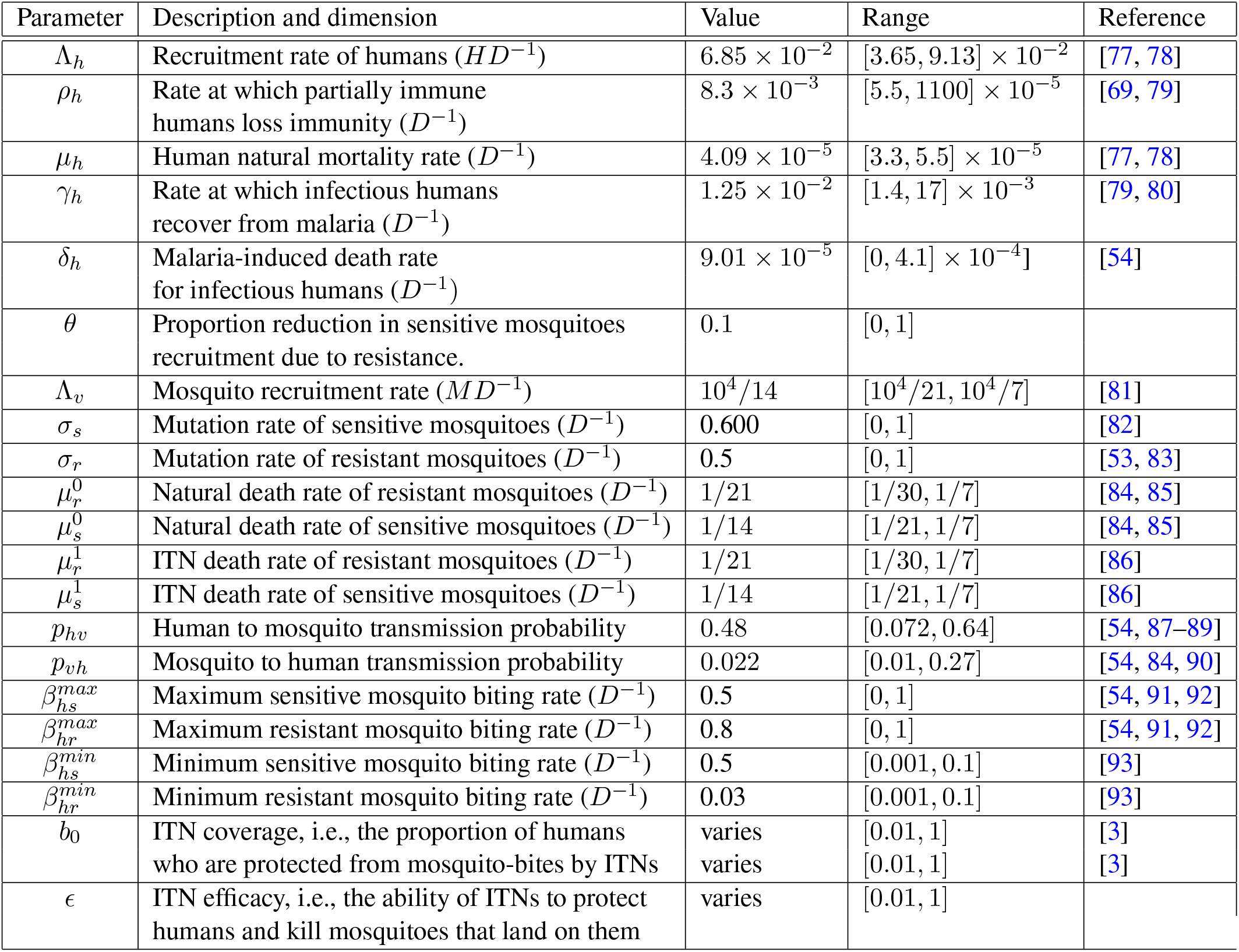
Table of parameter values and ranges used for the simulations of System (2.5). The dimension *H, D*, and *M* represent human, day, and mosquito, respectively. Dimensions are enclosed in parentheses at the end of parameter descriptions and excluded for dimensionless parameters.

where *A*_1_ = *δ*_*h*_ + *µ*_*h*_ + *γ*_*h*_, *A*_2_ = *µ*_*h*_ + *ρ*_*h*_, and the total human population is described by:

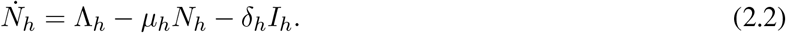

A Susceptible-infectious (SI) framework is used for the mosquito population. That is, the total mosquito population is broken down into susceptible and infectious mosquitoes. Furthermore, each of these two groups of mosquitoes is broken down into sensitive and resistant mosquitoes leading to the following compartmental classes: sensitive susceptible mosquitoes *S*_*s*_, which are susceptible mosquitoes that are not resistant to insecticides, resistant susceptible mosquitoes *S*_*r*_, which are mosquitoes that have developed resistance to insecticides, sensitive infectious mosquitoes *I*_*s*_, and resistant infectious mosquitoes, *I*_*r*_. It is assumed that mosquitoes can develop, as well as lose resistance to insecticides over time depending on the efficacy and coverage level of the treated bed nets. The development of resistance for sensitive mosquitoes occurs at per capita rate *σ*_*s*_, while the loss of resistance by resistant mosquitoes occurs at per capita rate *σ*_*r*_. Natural mortalities in the mosquito compartments occur at per capita rates 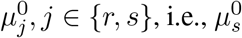 is the per capita rate of natural mortality in the sensitive mosquito compartments (*S*_*s*_ and *I*_*s*_) and 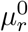 is the per capita rate of natural mortality in the resistant mosquito compartments (*S*_*r*_ and *I*_*r*_). Since ITNs are designed to prevent mosquitoes from biting humans who sleep under them and also to kill mosquitoes that land on them, we follow the approach in Ngonghala et al. [60, 69] and model the total mosquito mortality (i.e., natural and ITN-induced mortality) rate by the single term *µ*_*j*_, *j* ∈ {*r, s*} using the functional form 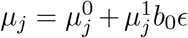, where 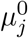 is the per capita natural mortality rate of mosquitoes in the sensitive classes (*j* = *s*) and resistant classes (*j* = *r*), 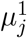 is the death rate of mosquitoes in the sensitive and resistant classes that land on ITNs, 0 ≤ *b*_0_ ≤ 1 is ITN coverage, and 0 ≤ *ϵ* ≤ 1 is the efficacy of ITNs. This implies that when ITN coverage and efficacy are high, mosquito mortalities resulting from ITN-use are also high and vice versa. In the context of this work, ITN-coverage refers to the proportion of humans who are protected by ITNs and ITN-efficacy refers to the ability of ITNs to protect humans who sleep under them from mosquito bites, as well as kill mosquitoes that land on them. The sensitive susceptible mosquito class is increased by births occurring at rate (1 − *θ*)Λ_*v*_, where 0 < *θ* < 1 is the proportion of mosquito births that are resistant and when resistant mosquitoes lose their resistance at per capita rate *σ*_*r*_. The population of this class reduces when sensitive susceptible mosquitoes become infected by infectious humans with force of infection *λ*_*hs*_, become resistant at per capita rate *σ*_*s*_, die naturally at per capita rate 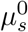, or die as a result of insecticides on nets at rate 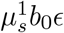. The sensitive infectious mosquito population is increased by incoming newly infectious sensitive mosquitoes *λ*_*hs*_*S*_*s*_ and is reduced by natural deaths occurring at rate 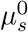, insecticide-induced deaths at rate 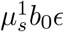, and when sensitive infectious mosquitoes become resistant at rate *σ*_*s*_. The populations of the resistant susceptible and infectious mosquitoes are increased or reduced through similar processes, with the subscript *s* in the parameters replaced by the subscript *r*. Using this description and the schematics in Fig. 1, the population and disease dynamics for the mosquitoes are described by the system of nonlinear first order ordinary differential equations:

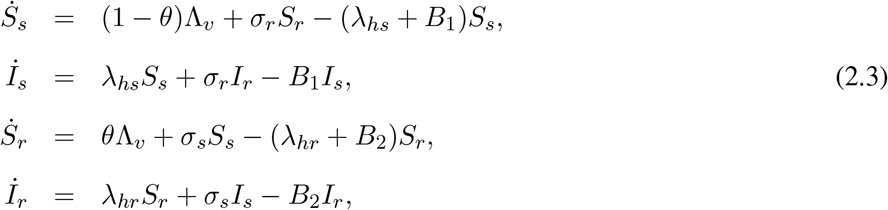

where *B*_1_ = *σ*_*s*_ + *µ*_*s*_, *B*_2_ = *σ*_*r*_ + *µ*_*r*_, and the dynamics of the total mosquito population is described by the equation

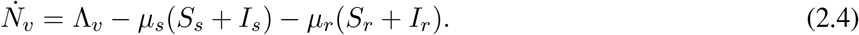

We differentiate the biting rate of mosquitoes such that sensitive and resistant mosquitoes bite humans at respective rates *β*_*hs*_ and *β*_*hr*_ [75, 76]. As in Ngonghala et al. [60, 69], we model these biting rates with the functional forms: 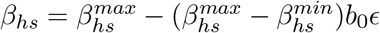 *and* 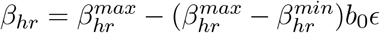. Observe that for each type of mosquito, the biting rate is maximum when *b*_0_ = 0 or *ϵ* = 0, i.e., when there is no ITN coverage or there is coverage with non-effective ITNs and minimum when *b*_0_ = 1 and *ϵ* = 1, i.e., when the entire population uses highly effective ITNs. The force of infection 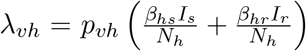, where *p*_*vh*_ is the probability that a bite from a sensitive or resistant infectious mosquito will infect a susceptible human. On the other hand, the forces of infection *λ*_*hs*_ and *λ*_*hr*_ are given by 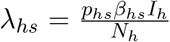 and 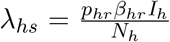, respectively. The full model that incorporates ITN-use, mosquito-resistance to insecticides, and differentiated infectivity captured through different biting rates by sensitive and resistant mosquitoes is described by the nonlinear system:

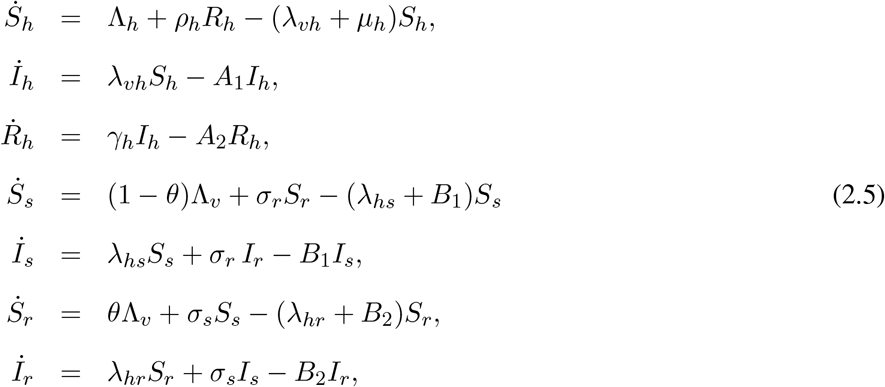

where the forces of infection *λ*_*vh*_, *λ*_*hs*_, and *λ*_*hr*_ are defined as:

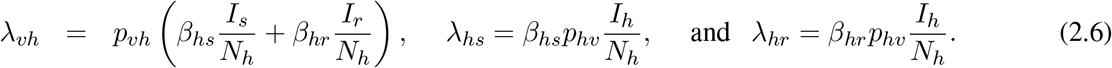

A summary of the description of model parameters together with baseline and ranges of numerical values for the parameters and their sources are presented in Table 1.

As human and mosquito populations, each of the variables *S*_*h*_, *I*_*h*_, *R*_*h*_, *S*_*s*_, *I*_*s*_, *S*_*r*_, and *I*_*r*_, and the parameters of the system (see Table 1) are non-negative. As in [60, 69], it is straight forward to verify that the system is well-posed within the epidemiologically feasible region

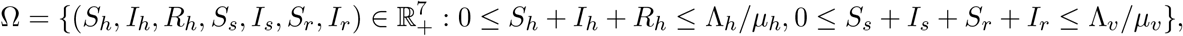

where *µ*_*v*_ = *min*(*µ*_*s*_, *µ*_*r*_). Considering the equation for total human population (2.2), we have: 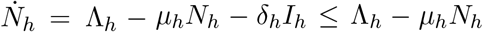. Therefore, 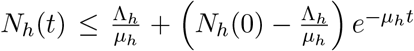, where *N*_*h*_(0) is the initial total human population. Similarly, if *µ*_*v*_ = *min*(*µ*_*s*_, *µ*_*r*_), then from the equation for the total mosquito population (Eq.(2.4)) becomes: 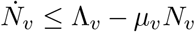. Thus, 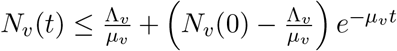, where *N*_*v*_(0) is the initial total human population. Observe that *N*_*h*_(*t*) ≤ Λ_*h*_*/µ*_*h*_ and *N*_*v*_(*t*) ≤ Λ_*v*_*/µ*_*v*_ as *t* → ∞. Therefore, the epidemiologically feasible region 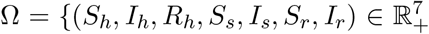 is positively invariant and attracting with respect to the model system (2.5). That is, any solution of the model with initial data within Ω is trapped within Ω for *t* > 0.

## 3. Model analysis

In this section, we determine the basic reproduction number and explore the existence and stability properties of equilibria to System (2.5). The basic reproduction number of the model system calculated using the next generation matrix approach [94] (see the online supplementary information for details) is

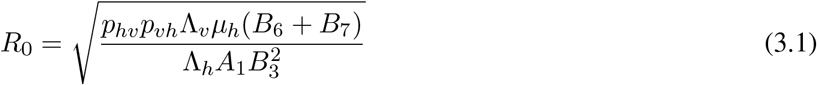

where

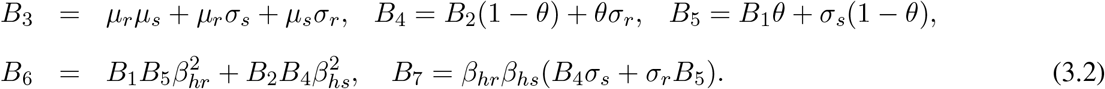

Equilibria are obtained by setting the left-hand side to zero and solving for the variables *S*_*h*_, *I*_*h*_, *R*_*h*_, *S*_*s*_, *I*_*s*_, *S*_*r*_, *I*_*r*_:

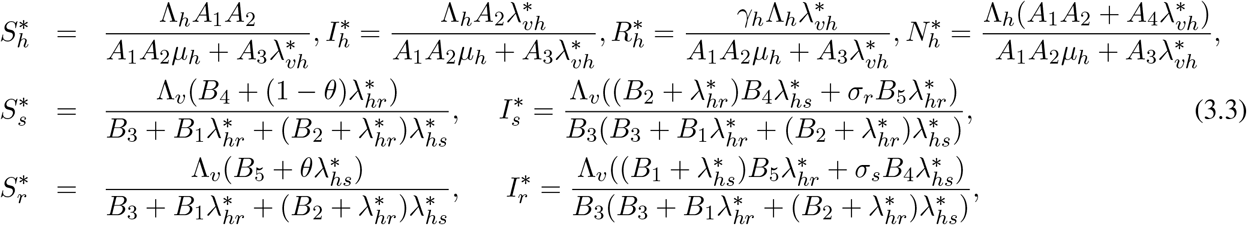

where *A*_3_ = *µ*_*h*_*A*_1_ + *ρ*_*h*_(*δ*_*h*_ + *µ*_*h*_) and *A*_4_ = *A*_2_ + *γ*_*h*_. Substituting 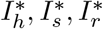, and 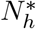 into the forces of infection *λ*_*hs*_, *λ*_*hr*_, and *λ*_*vh*_ from Eqs. (2.6) yields

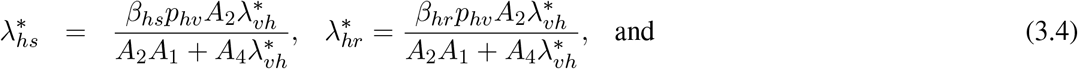

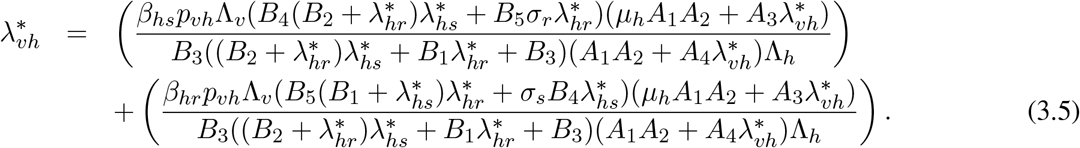

Substituting 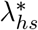 and 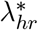 from Eqs. (3.4) into Eq. (3.5) and collecting terms in powers of 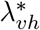 results in

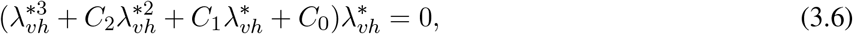

where

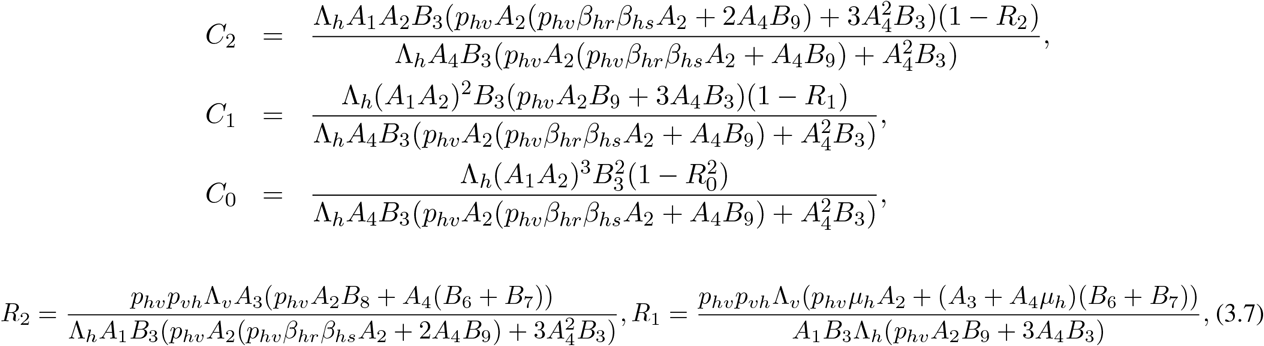

*B*_8_ = *β*_*hr*_*β*_*hs*_(*B*_4_*β*_*hs*_ + *B*_5_*β*_*hr*_), and *B*_9_ = *β*_*hr*_*B*_1_ + *β*_*hs*_*B*_2_. Note that *C*_0_ ≥ 0 if *R*_0_ ≤ 1, *C*_0_ < 0 if *R*_0_ > 1, *C*_1_ ≥ 0 if *R*_1_ ≤ 1, *C*_1_ < 0 if *R*_1_ > 1, and that *C*_2_ ≥ 0 if *R*_2_ ≤ 1, *C*_2_ < 0 if *R*_2_ > 1. Using Descartes’ rule of signs, we guess that Eqs. (2.5) can have zero, one, two, or three endemic equilibria, which leads to the Theorem:

#### Theorem 3.1.

*The model system* (2.5) *can have*

- *no endemic equilibrium point when C*_0_ ≥ 0, *C*_1_ ≥ 0, *and C*_2_ ≥ 0;
- *one possible endemic equilibrium point if C*_0_ < 0, *C*_1_ ≥ 0, *and C*_2_ ≥ 0, *or C*_0_ ≤ 0, *C*_1_ < 0, *and C*_2_ ≥ 0, *or C*_0_ ≤ 0, *C*_1_ ≤ 0, *and C*_2_ ≤ 0;
- *zero or two possible endemic equilibrium points if C*_0_ ≥ 0, *C*_1_ > 0 *and C*_2_ < 0, *or C*_0_ > 0, *C*_1_ < 0 *and C*_2_ ≥ 0, *or C*_0_ > 0, *C*_1_ ≤ 0 *and C*_2_ < 0.

#### Remark 3.2.

*The possibility of two endemic equilibrium points when C*_0_ ≥ 0, *which corresponds to R*_0_ ≤ 1 *postulated in Theorem 3.1 highlights the possibility of a backward (sub-critical bifurcation). We illustrate this using numerical simulations in Fig. 4. To identify specific conditions under which the polynomial equation* (3.6), *has a unique endemic equilibrium, or exactly two endemic equilibrium points, we can apply a combination of Sturm’s Theorem and Descartes rule of signs applied to a canonical form the equation obtained through the substitution* 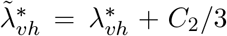 *(see [72] for details). An illustration of the case in which the model* (2) *has one endemic equilibrium is presented in Fig. 3*.

**Figure 3:**
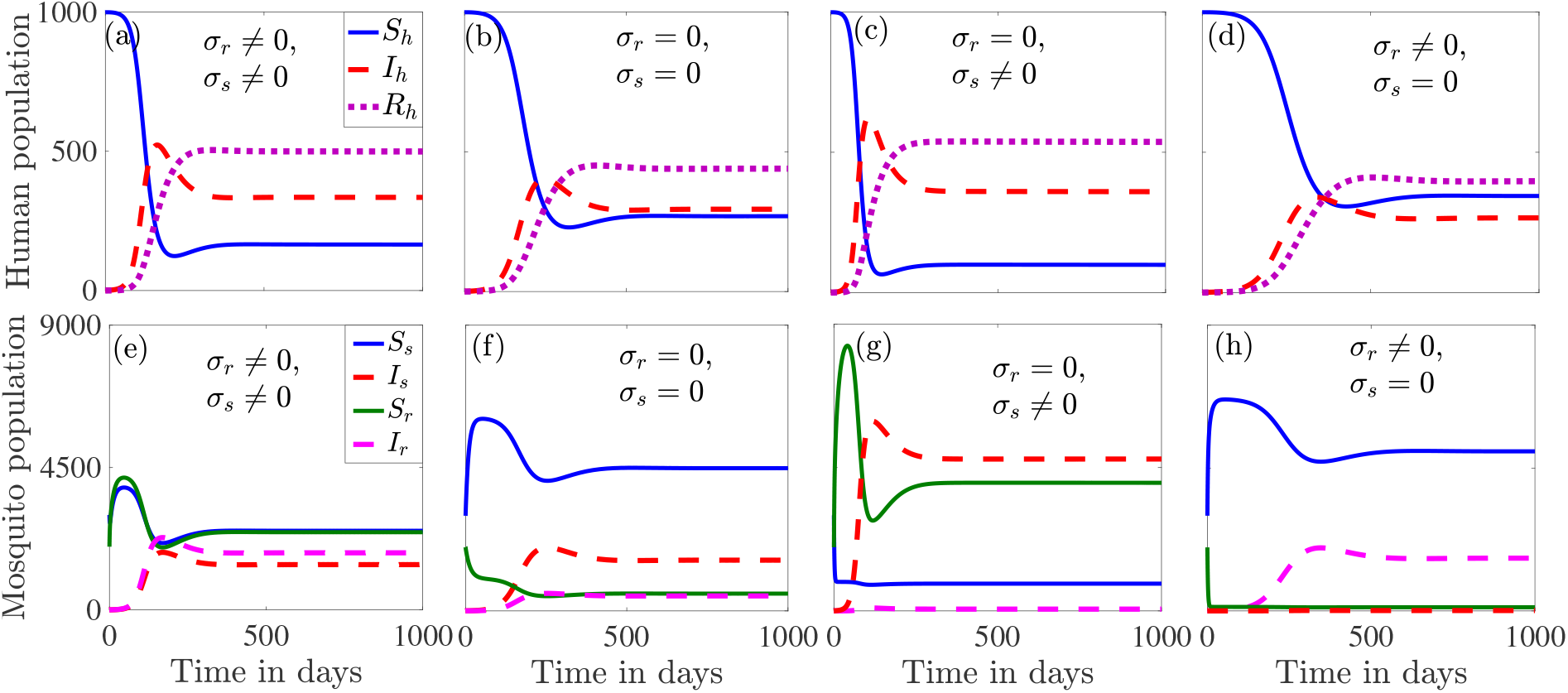
Numerical simulation results illustrating the existence of an endemic equilibrium to System (2) when *R*_0_ > 1. Graphs (a) and (e) show the dynamics for the case in which resistance is acquired at rate *σ*_*s*_ ≠ 0 and lost over time at rate *σ*_*r*_ ≠ 0. Graphs (b) and (f) illustrate the dynamics when resistance is acquired only through mosquito recruitment and is permanent. Graphs (c) and (g) illustrate the dynamical behavior of the system when resistance is permanent and can be acquired through mosquito recruitment or transition of sensitive mosquitoes to resistant mosquitoes at rate *σ*_*s*_ ≠ 0. Graphs (d) and (h) illustrate the dynamical behavior of the system when resistance is not permanent and can only be acquired through mosquito recruitment, i.e., *σ*_*r*_ ≠ 0 and *σ*_*s*_ = 0. Parameter values used for the simulations are presented in Table 1.

**Figure 4:**
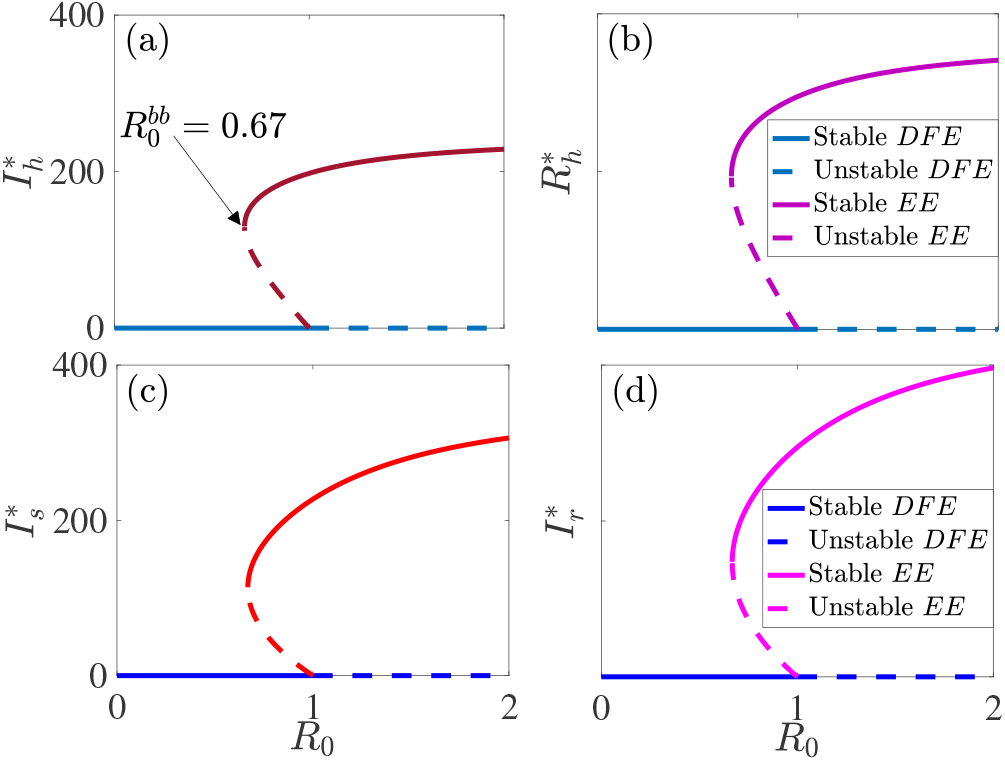
Bifurcation plot of the equlibrium infectious humans, 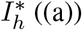, partially immune humans, 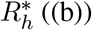 sensitive infectious mosquitoes, 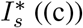, and the resistant infectious mosquitoes, 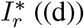 against the basic reproduction number, *R*_0_. Model (2) exhibits a backward bifurcation for 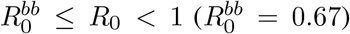, where an unstable endemic equilibrium (*EE*) is located between a stable endemic equilibrium (*EE*) and a stable disease free equilibrium (*DFE*). The ITN coverage (*b*_0_) is varied, while the other parameter values are as given in Table 1.

### 3.1. The disease-free equilibrium

The case of Eq. (3.6) for which 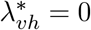 = 0 corresponds to the disease-free equilibrium 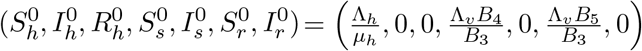. The Jacobian of System (2.5) evaluated at this disease-free equilibrium is:

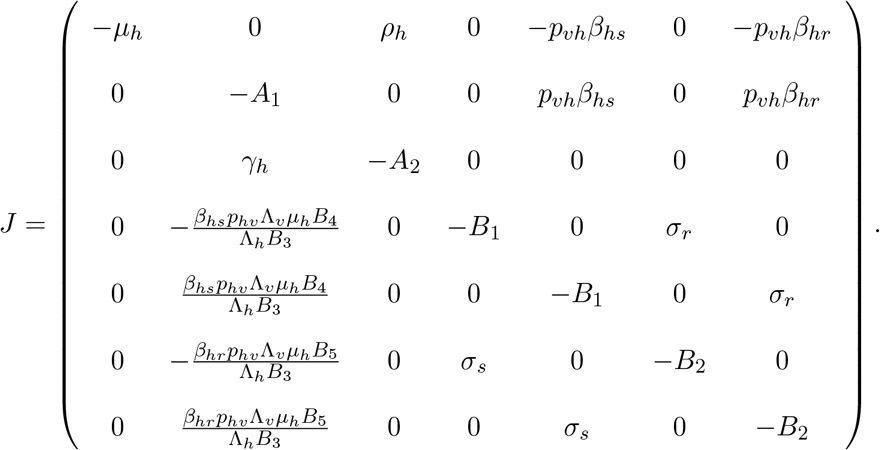

If *ξ* is an eigenvalue of *J*, then *ξ*_1_= − *µ*_*h*_, *ξ*_2_ = − *A*_2_, and 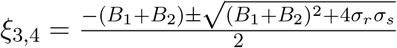 are four eigenvalues of *J*. Observe that both *ξ*_3_ and *ξ*_4_ are real and negative since 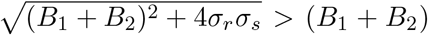. Therefore, four of the eigenvalues of *J* are real and negative. The other three eigenvalues are given by the cubic equation:

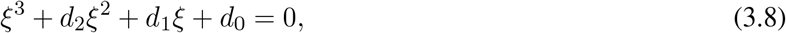

Where 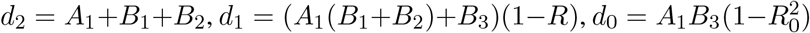, and 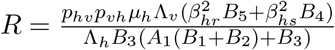 It can be verified that 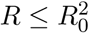. Hence, when *R*_0_ < 1, *d*_0_ > 0 and *d*_1_ > 0. Since *d*_1_ ∗ *d*_2_ − *d*_0_ > 0, the Routh-Hurwitz (2.5) is locally asymptotically stable when *R*_0_ < 1. condition assures us that no solution of the Eq. (3.8) is positive when *R*_0_ < 1. Therefore, all eigenvalues of *J* are negative or have negative real parts (if they are complex) when *R*_0_ < 1. This proves the following Theorem:

#### Theorem 3.3.

*The disease-free equilibrium* 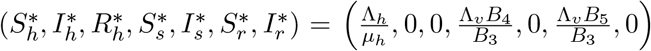 *of system (2.5) is locally asymptotically stable when R0 < 1*.

## 4 Results

The long-term dynamics of Model (2) illustrating the existence of a stable disease free equilibrium (*DFE*) when the basic reproduction number *R*_0_ is less than one and a stable endemic equilibrium when *R*_0_ > 1 are presented in Figs. 2-3. Parameters used for the simulations are presented in Table 1. For this parameter regime and with ITN coverage *b*_0_, chosen such that *R*_0_ < 1 (e.g., *b*_0_ = 90%), we obtain a disease free equilibrium depicted in Fig. 2. The case in which resistant mosquitoes can lose their resistance and sensitive mosquitoes develop resistance at recruitment and through mutation, i.e., *σ*_*r*_ ≠ 0, *σ*_*s*_ ≠ 0 is presented in Fig. 2 (a) and (c), while the case in which sensitive mosquitoes only develop resistance at recruitment and resistance is permanent, i.e., *σ*_*s*_ = 0, *σ*_*r*_ = 0 is presented in Fig. 2 (b) and (d).

For the parameter regime in Table 1, when resistant mosquitoes are able to lose resistance, i.e., *σ*_*s*_ ≠ 0, *σ*_*r*_ ≠ 0 or *σ*_*s*_ = 0, *σ*_*r*_ ≠ 0, there are more resistant infectious mosquitoes than sensitive infectious mosquitoes at equilibrium (Fig. 3 (e) and (h)). The disease is concentrated mostly among the sensitive mosquitoes when resistance is permanent, i.e., *σ*_*s*_ = 0, *σ*_*r*_ = 0 or *σ*_*s*_ ≠ 0, *σ*_*r*_ = 0 (Fig. 3 (f) and (g)). Additionally, the highest (respectively, lowest) disease prevalence is observed among the sensitive (respectively, resistant) mosquitoes when resistance acquired through mosquito-recruitment or transition from adult sensitive to resistant mosquito at per capita rate *σ*_*s*_ is permanent (Fig. 3 (g)). On the other hand if sensitive mosquitoes only become resistant through mosquito recruitment, i.e., *σ*_*s*_ = 0 and resistance is not permanent, i.e., *σ*_*r*_ ≠ 0, then the resistant mosquito population only exists at very low levels, while disease prevalence is predominantly among the human and sensitive mosquito population.

Figure 4 shows that Model (2) exhibits a backward bifurcation when *R*_0_ < 1. In this case, a stable disease free equilibrium (*DFE*) co-exists with a stable endemic equilibrium (*EE*) for 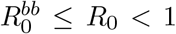, where the backward bifurcation threshold, 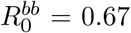. Observe that when 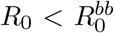, we have only a stable disease free equilibrium. Thus, disease control measures must be applied continuously to reduce *R*_0_ below 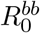.

Next, we identify important model parameters that drive the system and that can be targeted for control purposes through a global uncertainty and sensitivity analyses using the Latin-Hypercube Sampling (LHS) and Partial Rank Correlation Coefficient (PRCC) methods (see the online supplementary information for details on the methodology). We found out that uncertainty or variability in the efficacy of ITNs *ϵ*, and ITN coverage *b*_0_, contribute most to uncertainty or variability in the basic reproduction number *R*_0_, and the threshold parameter groupings *R*_1_ and *R*_2_ from Eq. (3.7) (Fig. 5). Each of these thresholds is useful for determining the existence of endemic equilibria and hence disease prevalence or disease elimination. There is a negative correlation between each of these thresholds and ITN efficacy and coverage, i.e., raising ITN efficacy or coverage results in a reduction in the basic reproduction number, *R*_1_, and *R*_2_. Thus, allowing these parameters to fall will trigger an increase in the basic reproduction number, *R*_1_, and *R*_2_, and hence an increase in disease prevalence. Other important parameters that can be targeted for control include the maximum biting rate of resistant mosquitoes 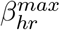, the transmission probability from infectious humans to sensitive and resistant susceptible mosquitoes *p*_*hv*_, the transmission probability from sensitive and resistant mosquitoes to susceptible humans *p*_*vh*_, and the mosquito recruitment rate Λ_*v*_. Other important parameters that do not impose as much variability and uncertainty as the above are the maximum biting rate of sensitive mosquitoes *βhs*^*max*^ and the natural mortality rates of sensitive and resistant mosquitoes. Among the least influential parameters are the minimum biting rates of sensitive and resistant mosquitoes *βhs*^*min*^ and *βhr*^*min*^, respectively, confirming the low malaria risk in areas with low mosquito densities, or where mosquitoes are not allowed to bite humans. It is worth mentioning that although the human recruitment rate Λ_*h*_, has a high PRCC, we have not highlighted it here since we are interested only in parameters that can used for malaria control purposes.

**Figure 5:**
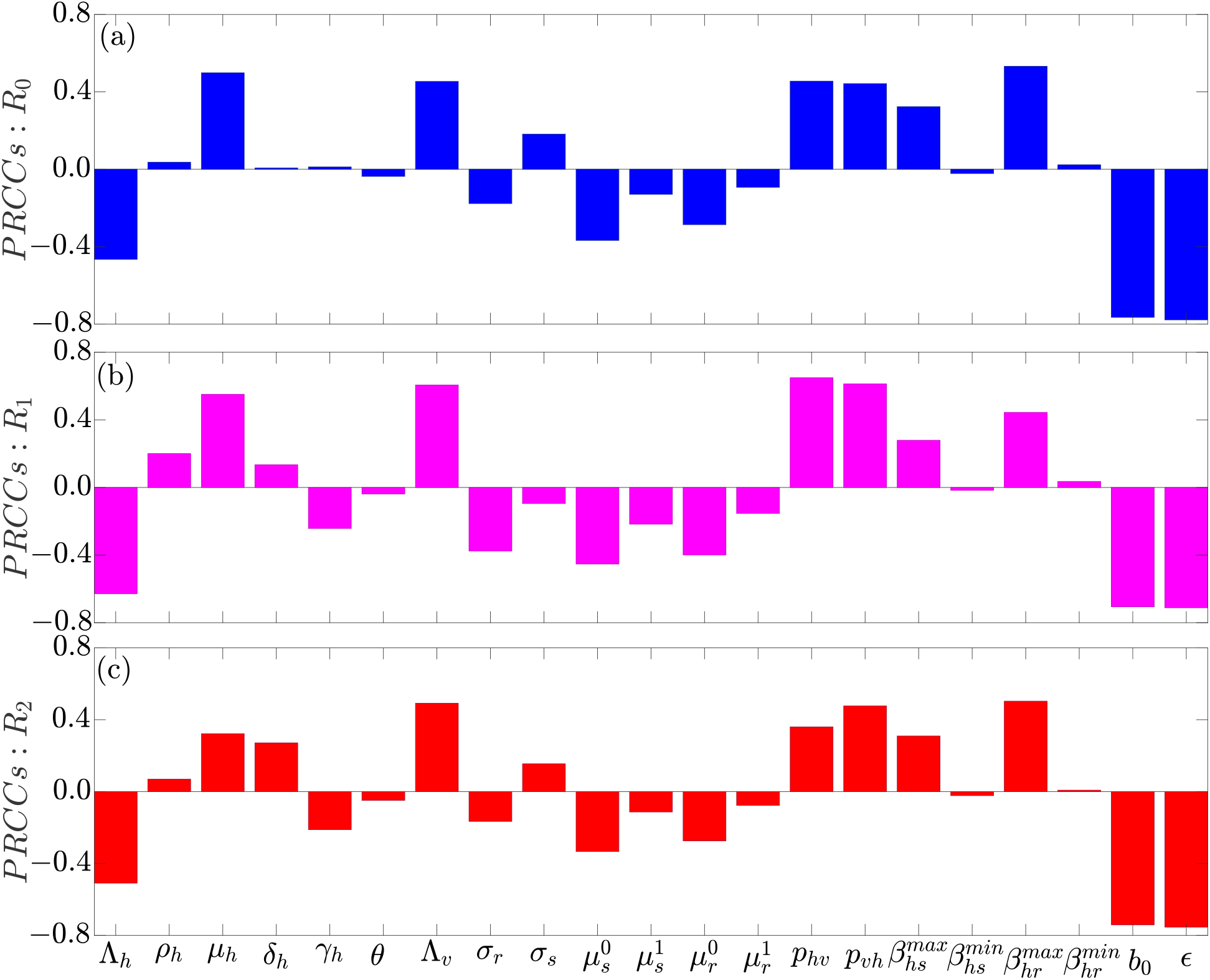
Global uncertainty and sensitivity analysis results showing the contributions of uncertainty or variability in model parameters to uncertainty or variability to the (a) basic reproduction number *R*_0_, and (b)-(c) threshold quantities *R*_1_ and *R*_2_ (Eq. (3.7)). Positive PRCCs represent positive correlation, i.e., an increase in a parameter will trigger an increase in *R*_0_, *R*_1_, or *R*_2_, while negative PRCCs represent negative correlations. The magnitude of the PRCC represents the level of significance. Parameters used for the simulations are presented in Table 1.

Also, our numerical results indicate that uncertainty or variability in the efficacy of ITNs *ϵ*, ITN coverage *b*_0_, the human recovery rate from infection *γ*_*h*_, the maximum biting rate of resistant mosquitoes 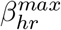, the transmission probability from infectious humans to infectious mosquitoes *p*_*hv*_, and the mosquito recruitment rate Λ_*v*_, are more influential in imposing variability or uncertainty to the infectious human, resistant, and sensitive mosquito populations (Fig. 6).

**Figure 6:**
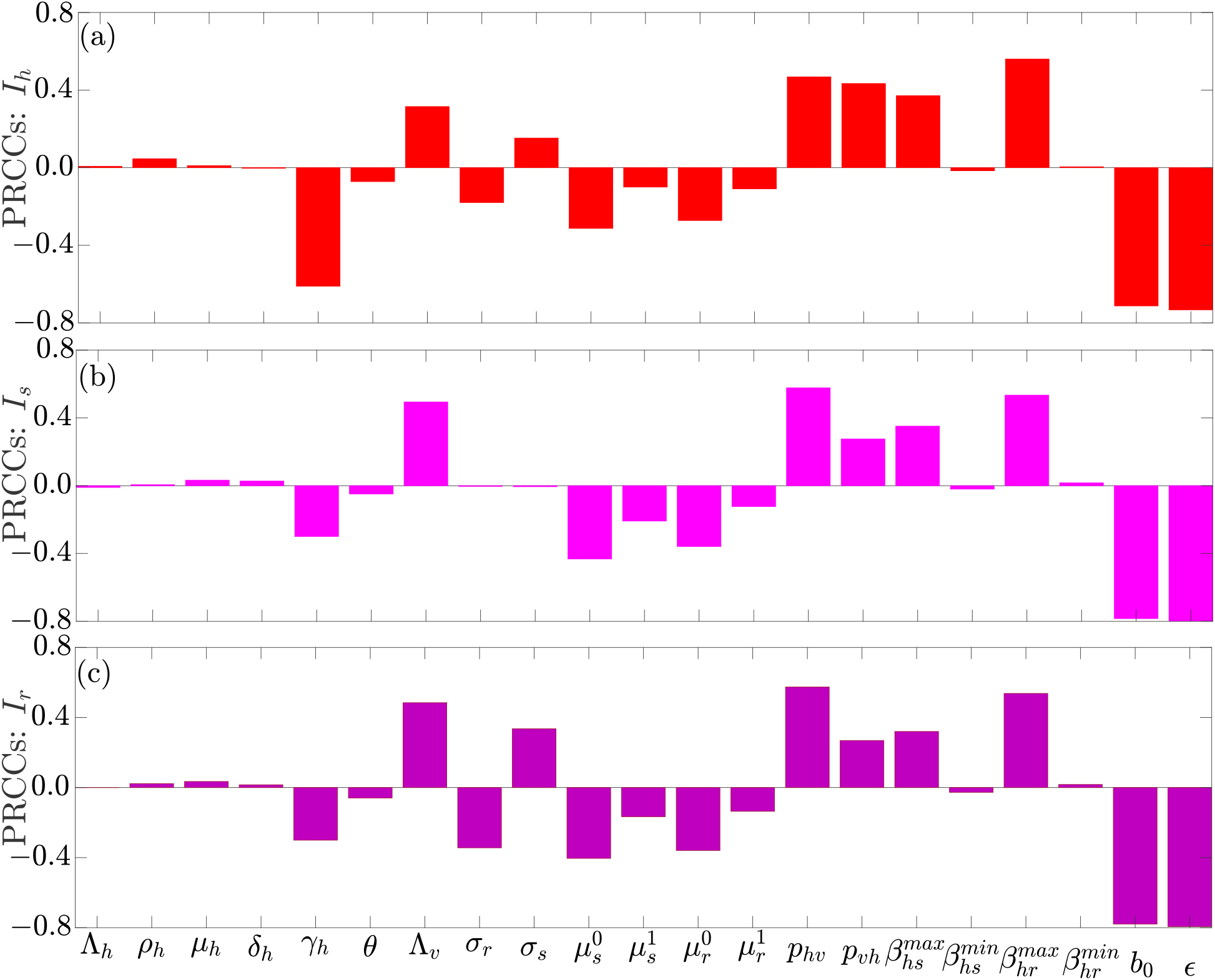
Global uncertainty and sensitivity analysis results showing the contributions of uncertainty or variability in model parameters to uncertainty or variability to the (a) infectious human population (*I*_*h*_), (b) sensitive mosquito population (*I*_*s*_), and (c) resistant mosquito population (*I*_*r*_). Positive PRCCs represent positive correlations, i.e., an increase in a parameter will trigger an increase in *I*_*h*_, *I*_*s*_, or *I*_*r*_, while negative PRCCs represent negative correlations. The magnitude of the PRCC represents the strength or level of significance. Parameters used for the simulations are presented in Table 1.

Next, we investigate the impact of ITN coverage (*b*_0_), ITN efficacy (*ϵ*), and resistance to insecticide used in ITNs by mosquitoes through the resistance acquisition and loss rates *σ*_*s*_ and *σ*_*r*_, respectively, on a key measure of disease intensity–the basic reproduction number. Figure 7 shows ITN coverage levels for different ITN efficacy and acquisition and loss rates of resistance appropriate for containing the malaria disease. For very high ITN efficacy, e.g., 100%, approximately 76% ITN coverage is necessary for containing the disease, while for ITN efficacy below 76%, even 100% ITN coverage is not enough for containing the disease (Fig. 7(a)). It is worth noting that the 76% mentioned here is the horizontal (*b*_0_) axis coordinate of the point of intersection of the dotted green curve and the line *R*_0_ = 1. Other ITN coverage levels required to contain the disease under different ITN efficacy are obtained in a similar way using appropriate curves that correspond to the chosen level of efficacy.

**Figure 7:**
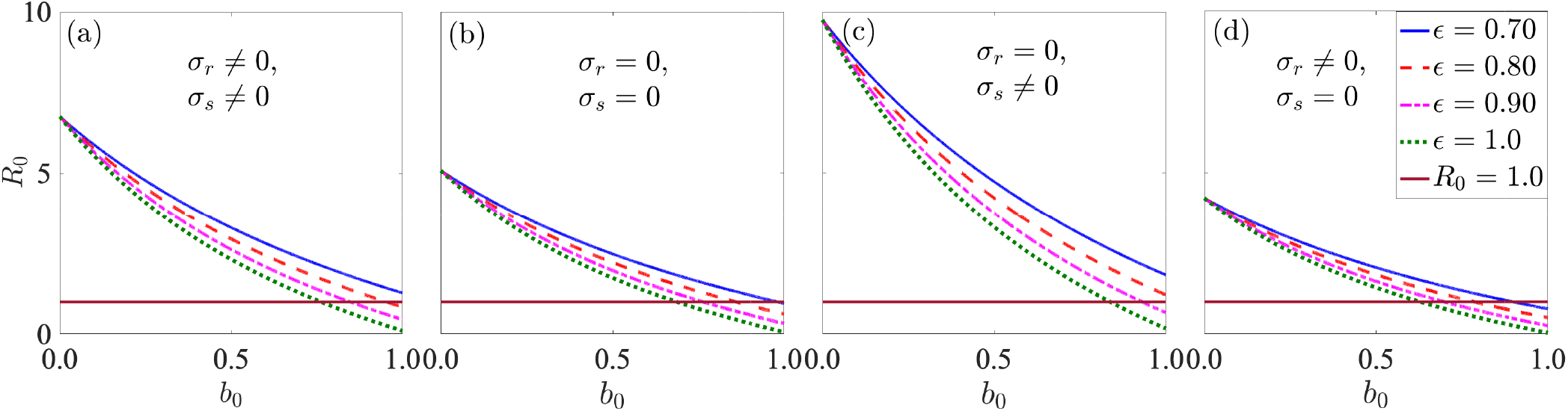
Numerical simulations of the basic reproduction number *R*_0_, against ITN coverage *b*_0_, illustrating insecticide-impregnated bed-net coverage levels required for bringing malaria under control for four values of insecticide-impregnated bed-net efficacy (*ϵ*) when mosquito resistant to insecticides is permanent or nonpermanent. (a) Mosquito resistance to insecticides is not permanent and acquired through mosquito resistant and through transition of mosquitoes occurring at rate *σ*_*s*_ ≠ 0. (b) Mosquito resistance is permanent and is only acquired through mosquito recruitment. (c) Resistance is permanent and acquired through mosquito recruitment and adult sensitive mosquito transition. (d) Resistance is not permanent and is acquired only through mosquito-recruitment. Other parameters used for the simulations are presented in Table 1.

Figures 7(a) and (d) correspond to behavioral resistance to insecticides, which is not permanent. Unlike behavioral resistance where resistant mosquitoes lose their resistance over time, resistance to insecticides can be permanent. That is once a mosquito becomes resistant, it maintains this status until death. This is the case with metabolic and cuticle resistance [41–43]. Hence, we consider three slightly simplified versions of the model: 1) The case in which there is no transition between the sensitive and resistant mosquito classes (*σ*_*r*_ = *σ*_*s*_ = 0). That is, we are assuming that resistance is acquired only through mosquito-recruitment and is permanent (Fig. 7(b)). 2) The case in which resistance is permanent and acquired both through mosquito recruitment and adult sensitive mosquitoes becoming resistant, i.e., *σ*_*r*_ = 0, *σ*_*s*_ ≠ 0 (Fig. 7(c)). 3) The case in which resistance is not permanent and acquired only through mosquito recruitment, i.e., *σ*_*r*_ ≠ 0, *σ*_*s*_ = 0 (Fig. 7(d)). For the case in which resistance is permanent and acquired only through mosquito recruitment (Fig. 7(b)), when ITN efficacy is 70%, 80%, 90%, or 100% approximately 98%, 86%, 76% or 69% ITN coverage, respectively, is required to contain the disease. However, for ITN efficacy below 68% even full ITN coverage might not be enough for containment. For the case in which resistance is permanent and acquired both through mosquito recruitment and transition of mosquitoes from the sensitive to the resistant compartmental class (Fig. 7(c)), when ITN efficacy is 90%, or 100% approximately 93% or 84% ITN coverage, respectively, is required to contain the disease. However, for ITN efficacies below 84% even full ITN coverage might not be enough for containment. For the case in which resistance is not permanent and only through mosquito recruitment (Fig. 7(d)), when ITN efficacy is 70%, 80%, 90%, or 100% about 90%, 79%, 70%, or 63% ITN coverage, respectively, is required to contain the disease. But for ITN efficacies below 63% even full ITN coverage might not be enough for containment.

Furthermore, we investigate the ITN coverage levels required for disease containment for different maximum mosquito biting rates, 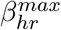and 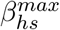 (Fig. 8(a) and (b)), ITN-induced mosquito mortality rates, 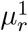 and 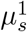, (Fig. 8(c) and (d)) and resistance acquisition and loss rates, *σ*_*s*_ and *σ*_*r*_, respectively, (Fig. 8(e) and (f)). When either the sensitive or resistant mosquitoes bite a lot, even 100% ITN coverage might not be enough to contain malaria. However, when either sensitive or resistant mosquitoes do not bite a lot, there is a threshold level of ITN coverage that might be enough to contain the disease. For example, if sensitive mosquitoes do not bite, while the biting rate of resistant mosquitoes is 0.5 per day, about 75% ITN coverage with efficacy of 90% is required to contain the disease (Fig. 8 (b)). Insecticide treated nets must be complemented with other control measures if their efficacy is below 70%.

**Figure 8:**
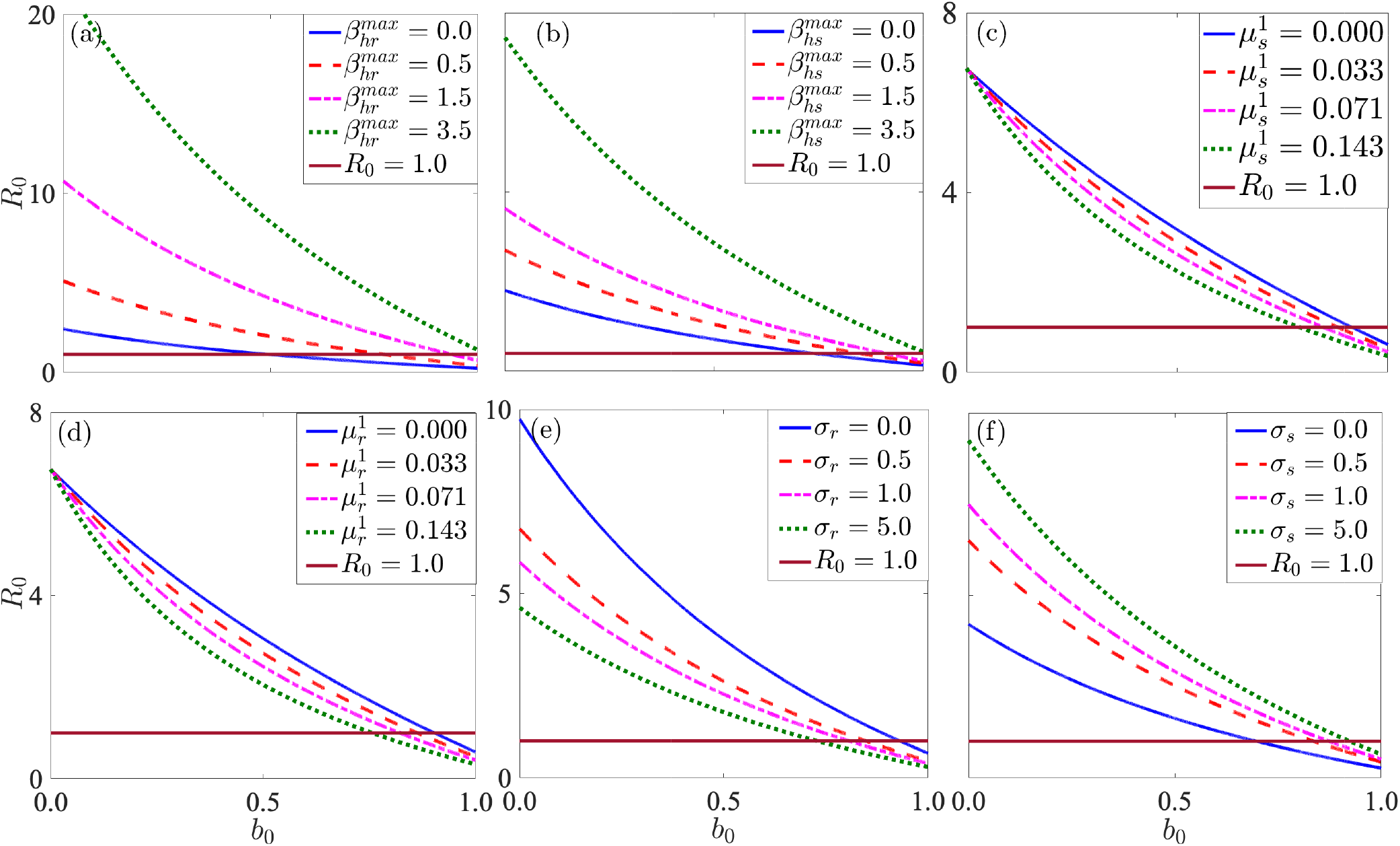
Numerical simulation results of the basic reproduction number *R*_0_, against ITN coverage *b*_0_, illustrating various insecticide-impregnated bed-net coverage levels required for bringing malaria under control for four values of (a)-(b) the maximaum biting rates of resistant and sensitive mosquitoes (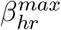and 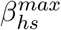,respectively), (c)- (d) insecticide-induced mortality rates of resistant and sensitive mosquitoes (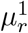 and 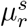),and (e)-(f) the resistant development and loss rates for mosquitoes (*σ*_*r*_ and *σ*_*s*_, respectively). Other parameters used for the simulations are presented in Table 1.

Our analysis also shows that if the sensitive mosquitoes do not bite humans or have a very low biting rate, a higher ITN coverage level is required than when the biting rate of resistant mosquitoes is low. For example, when ITN efficacy is 90% and the biting rate of sensitive mosquitoes is 0.0 or 0.5 per day, about 74% or 85% ITN coverage, respectively, is required to contain malaria (Fig. 8 (b)), while if the biting rate of resistant mosquitoes is 0 or 0.5 per day, approximately 50% or 77% ITN coverage, respectively, is required to get rid of malaria (Fig. 8 (a)). On the other hand, when ITNs do not kill sensitive or resistant mosquitoes that land on them, over 90% ITN coverage is required for containing malaria (Fig. 8 (c) and (d)). However, when ITNs kill mosquitoes that land on them, over 82% ITN coverage is required for eradication if the respective ITN killing rates for sensitive and resistant mosquitoes are 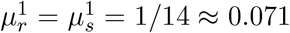. If the killing ability of ITNs is stronger, e.g., 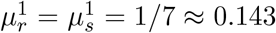, then less ITN coverage (approximately 75%) is required for malaria containment. When resistance to insecticides is permanent, about 93% ITN coverage is required (Fig. 8 (e)). However, when resistant mosquitoes lose their resistance over time at respective rates 0.5, 1.0, or 5.0 per day, approximately 85%, 81%, or 73% ITN coverage is required for eliminating malaria (Fig. 8 (e)). When sensitive mosquitoes become resistant at respective rates 0.0, 0.5, 1.0, or 5.0 per day, about 70%, 84%, 87%, or 92% ITN coverage is required for eliminating malaria (Fig. 8 (f)).

In the next set of results (Figs. 9-11), we present heat maps to demonstrate the impact of ITN coverage and one other parameter, e.g., ITN efficacy, maximum biting rate of mosquitoes, development and loss rates of resistance, etc., on two measures of disease intensity–the basic reproduction number *R*_0_ and the equilibrium infectious human populations, 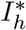. Similar results for the sensitive and resistant infectious mosquito populations and for the threshold parameters *R*_1_ and *R*_2_ are presented in the online supplementary information (SI).

**Figure 9:**
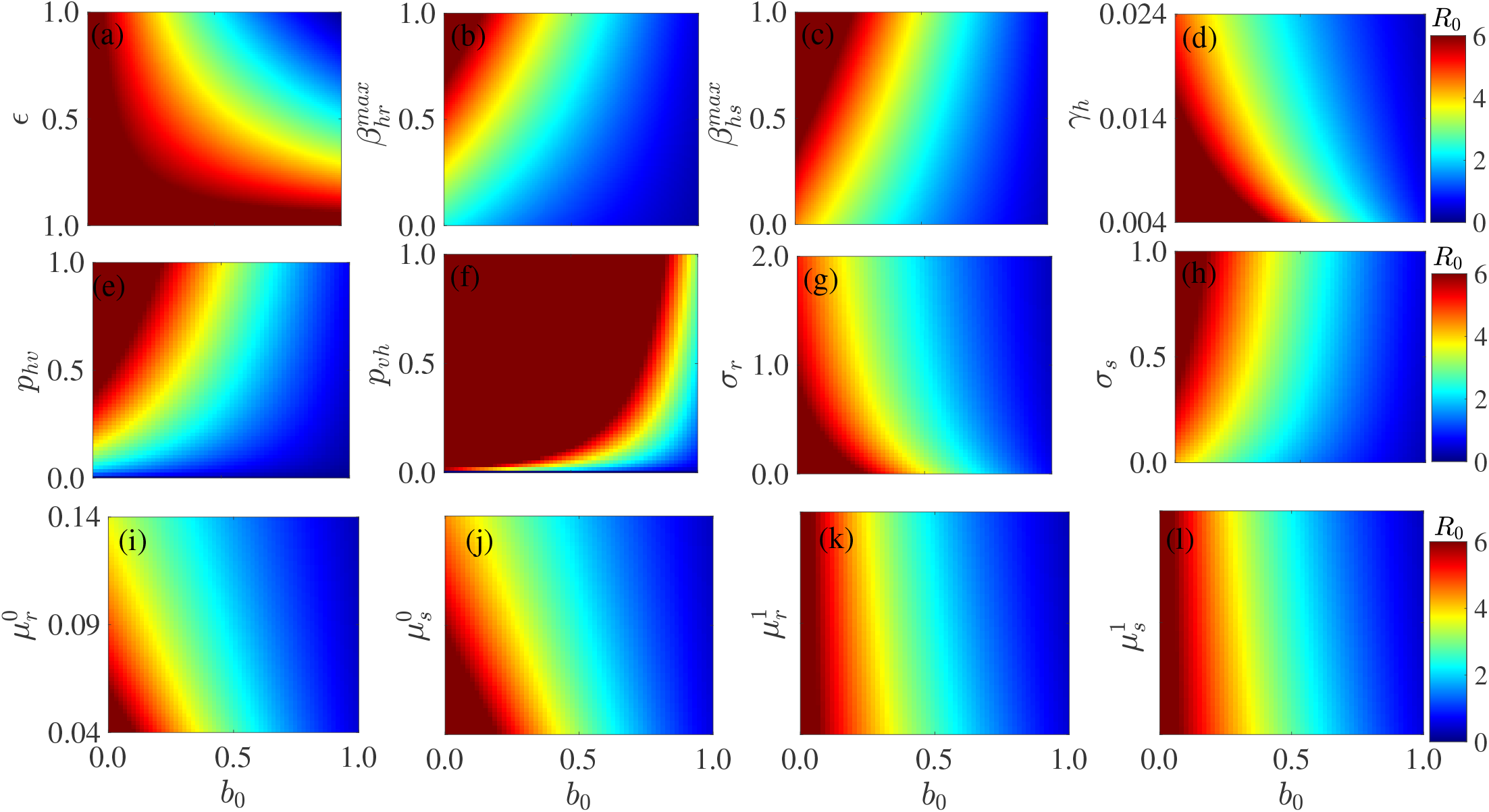
Heat maps from numerical simulations illustrating the impact on the basic reproduction number *R*_0_, of ITN coverage *b*_0_ and (a) ITN efficacy *ϵ*, (b) the maximum biting rate of resistant mosquitoes 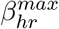, (c) the maximum biting rate of sensitive mosquitoes 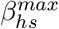, (d) the recovery rate from infection *γ*_*h*_, (e) the probability of an infectious human infecting a susceptible mosquito *p*_*hv*_, (f) the probability of infectious mosquito infecting a susceptible human *p*_*vh*_, (g) the rate at which resistant mosquitoes lose resistance *σ*_*r*_, (h) the rate at which sensitive mosquitoes develop resistance *σ*_*s*_, (i) the natural mortality rate of resistant mosquitoes 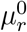, (j) the natural mortality rate of sensitive mosquitoes 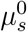, (k) the ITN-induced mortality rate of resistant mosquitoes 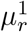, and (l) the ITN-induced mortality rate of sensitive mosquitoes 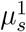. The values of the other parameters are presented in Table 1.

Disease prevalence is highest in areas in which fewer people are protected by ITNs when the efficacy of ITNs is very low (Fig. 9(a) and Fig. 10(a)). As observed earlier, when ITN efficacy is low, it becomes difficult to contain the disease even if everybody uses ITNs for protection and vice versa. There will also be more infectious individuals in the population when fewer humans are protected by ITNs and the human recovery rate from infection, the rate at which resistant mosquitoes lose resistance, or the rate at which mosquitoes die (naturally or as a result of ITN-use) is low, (Fig. 9(d), (g), (i)-(l) and Fig. 10(d), (g), (i)-(l)). On the other hand, disease prevalence is highest for combinations of low ITN coverage and high mosquito biting rate, high probability of humans infecting mosquitoes, when more sensitive mosquitoes develop resistance (Fig. 9(b), (c), (e), (h) and Fig. 10(b), (c), (e), (h)).

**Figure 10:**
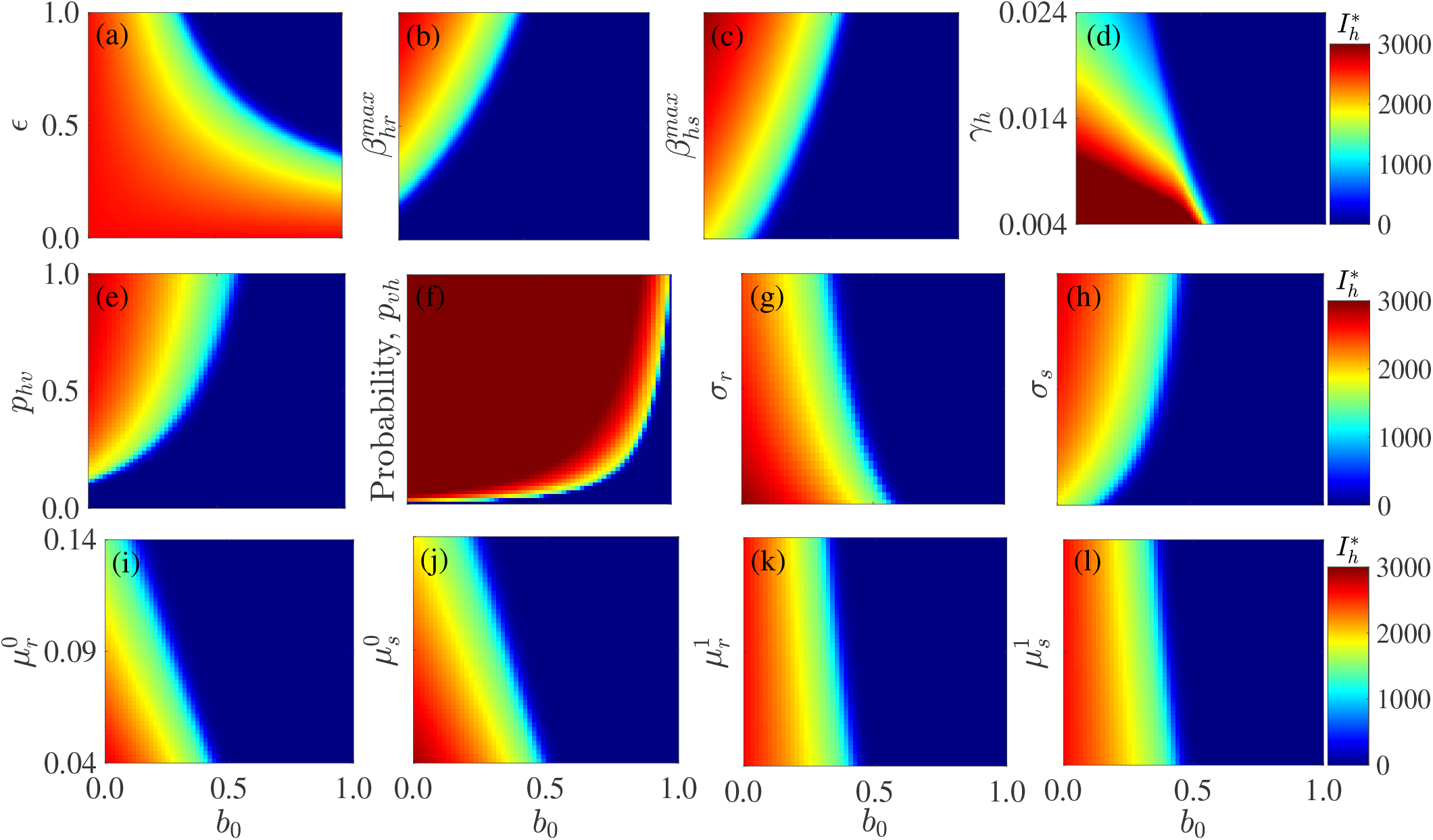
Numerical simulation results illustrating the impact on the Infectious human population *I*_*h*_, of ITN coverage *b*_0_ and (a) ITN efficacy *ϵ*, (b) the maximum biting rate of resistant mosquitoes 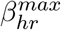, (c) the maximum biting rate of sensitive mosquitoes 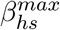, (d) the recovery rate from infection *γ*_*h*_, (e) the probability of an infectious human infecting a susceptible mosquito *p*_*hv*_, (f) the probability of infectious mosquito infecting a susceptible human *p*_*vh*_, (g) the rate at which resistant mosquitoes lose resistance *σ*_*r*_, (h) the rate at which sensitive mosquitoes develop resistance *σ*_*s*_, (i) the natural mortality rate of resistant mosquitoes 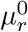, (j) the natural mortality rate of sensitive mosquitoes 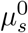, (k) the ITN-induced mortality rate of resistant mosquitoes 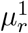, and (l) the ITN-induced mortality rate of sensitive mosquitoes 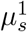. Other parameters used for the simulations are presented in Table 1.

Figure 11 shows heat maps of the impact on the basic reproduction number (Fig. 11 (a)-(c)), the infectious human population (Fig. 11 (d)-(f)), and the resistant infectious mosquito population (Fig. 11 (g)-(i)) for combinations of the maximum biting rate of resistant mosquitoes and the development rate of resistance, the loss rate of resistance, and the human recovery rate from infection. Disease prevalence is reduced, i.e., disease control is more feasible in areas of low resistant mosquito populations or when resistant mosquitoes do not bite a lot (Fig. 11 (a), (d), and (g)). Disease control is also feasible when more resistant mosquitoes lose their resistance or more humans recover fast from infection. On the other hand, disease prevalence is high in areas with high resistant mosquito densities (or when resistant mosquitoes bite more) and when more sensitive mosquitoes develop resistance.

**Figure 11:**
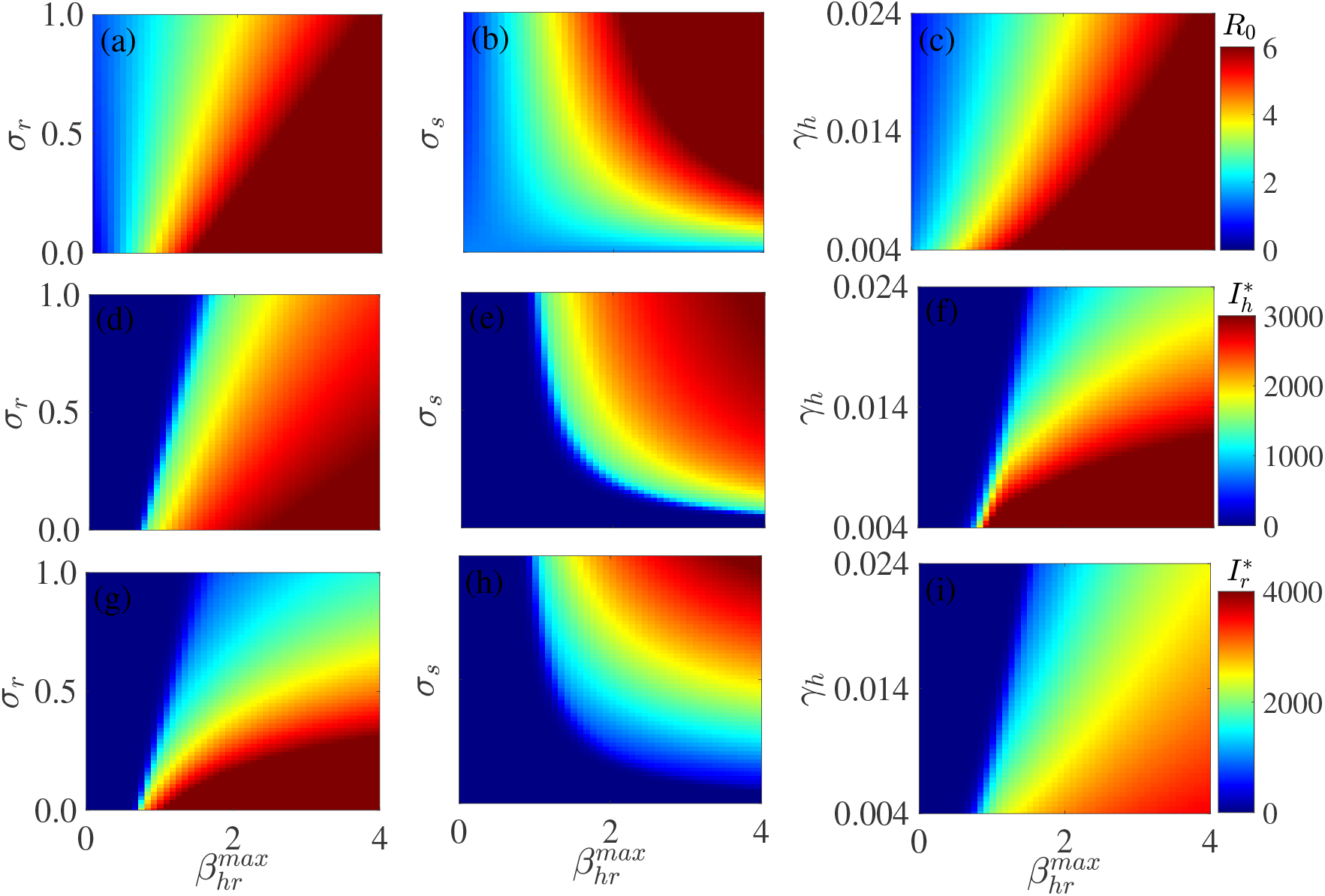
Numerical simulation results illustrating the effects on the basic reproduction number ((a)-(c)), the infectious human population ((d)-(f)), and the resistant infectious mosquito population ((g)-(i)) for combinations of the maximum biting rate of resistant mosquitoes 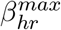 the rate at which resistance is lost, *σ*_*r*_ ((a), (d), (g)), and the development rate of resistance, *σ*_*r*_ ((b), (e), (h)), and the human recovery rate, *σ*_*r*_ ((c), (f), (i)). The other parameter values used for the simulations are presented in Table 1.

## 5. Conclusion

Malaria prevalence in sub-Saharan Africa remains high, despite the tremendous success in control efforts recorded over the past decade. For example, although some counties in Kenya boast of up to 80% personal protection through ITNs [95], malaria is still a major problem to the country. The gains of malaria control programs, especially those related to vector control such as ITNs and IRS continue to be dampened by human behavior, natural deterioration in ITN efficacy, misuse, and resistance to insecticides developed by mosquitoes. In this study, we developed and used a compartmental model to explore the interplay between ITN coverage, ITN efficacy, and resistance exhibited by mosquitoes to insecticides in relation to malaria prevalence and control.

Our results indicate that ITN efficacy and coverage are very important parameters to pay attention to in the fight against malaria. We found out that low ITN efficacy, or differentiated adherence to the use of ITNs has a negative impact on the outcomes of malaria risk and control. We also found out that as long as mosquitoes are resistant to insecticides, a combination of low ITN efficacy and high coverage, or high ITN efficacy and low coverage is not enough for reducing malaria to appreciable levels. The situation is worst when resistance to insecticides is permanent, i.e., for the case of metabolic or cuticle resistance. Hence, disease containment and possible elimination might be impossible when either ITN coverage or ITN efficacy is low and ITNs are not complemented with other control measures such as IRS, treatment, eliminating mosquito breeding sites near homes, etc. This is consistent with empirical studies in Ref. [15] indicating that both ITN efficacy and coverage for at risk populations must be high in order to achieve the target reduction in malaria prevalence. However, our results indicate that high ITN efficacy and moderately high ITN coverage and vice versa can be enough for appreciable reduction in malaria prevalence under certain circumstances, e.g., when resistance is only through mosquito recruitment (either permanent or not) or when more resistant mosquitoes are killed by ITNs. Consistent with common practice and public health recommendations, our results indicate that reducing mosquito populations, e.g., through killing, or eliminating their breeding sites near human homes and preventing mosquito bites, especially in areas of high mosquito density and high malaria prevalence are important for disease control. The fact that the model (2) exhibits a backward bifurcation implies that the disease can no longer be contained just by bringing the basic reproduction number *R*_0_, slightly below one. Instead, more and sustained control measures to reduce *R*_0_ below the new threshold value 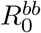, are required.

Finally, our analysis and results indicate that reducing resistance to insecticides is an important step towards malaria elimination. In fact, the 1:1.6 optimal target for containing malaria, which in itself is a challenge to attain [20] underestimates the effort required to contain malaria, especially in the presence of resistance. With this coverage level, elimination is impossible even when ITN efficacy is very high, e.g., 90-100%, unless when resistant mosquitoes do not bite, which at the moment is an impossibility. Therefore, designing control measures that prevent the development of resistance, or that target and eliminate resistant mosquitoes will improve disease control. This might involve using chemicals that mosquitoes might not easily resist or switching to new chemicals that mosquitoes are not resistant to for treating both long lasting and regular bed-nets.

Limitations of the current study involve the assumptions that ITN efficacy over the useful life of ITNs as prescribed by the World Health Organization (three years) and mosquito resistance to insecticides are both constant. However, these quantities might change over time, with ITN efficacy waning and resistance to insecticides strengthening. In fact, the development of resistance occurs over time with the frequency of new resistant vectors increasing with each generation. These limitations and other aspects of the malaria disease are currently under investigation and will be reported in a separate paper.

## Data Availability

The codes used for the simulations are available upon request

## Acknowledgement

CNN acknowledges the support of the Simons Foundation (Award #627346). CNN and JK thank the Department of Mathematics, University of Florida for partially funding JK’s visit to the Department and for providing office space and other resources necessary for collaborating on this project. JK acknowledges support from EMS-Simons for Africa, IMU-Simons African Fellowship Program and the School of Mathematics, University of Nairobi, for providing the necessary support and making it possible for JK to visit and collaborate on this project at the University of Florida.

## References

[1] Centers for Disease Control and Prevention, Malaria, Technical Report, Assessed on February 29, 2020.

[2] World Health Organization, World malaria report 2019, World Health Organization (Accessed on March 10, 2019).

[3] WHO, World malaria report 2018 WHO World Malaria Report, World Health Organization, Geneva., 2018.

[4] A. W. Sabrina, P. E. Kaufman, CDC 2010, African malaria mosquito Anopheles gambiae Giles (Insecta: Diptera: Culicidae), URL: https://edis.ifas.ufl.edu/in1048.

[5] J. Hemingway, N. Hawkes, L. McCarroll, H. Ranson, The molecular basis of insecticide resistance in mosquitoes., Insect Biochemistry and Molecular Biology 34 (2004) 653–665.

[6] Ministry of Health, The epidemiology and control profile of malaria in Kenya, National Malaria Control Programme (2016).

[7] W. Takken, W. Snellen, J. Verhave, B. Knols, S. Atmosoedjono, Environmental measures for malaria control in Indonesia: an historical review on species sanitation., Wageningen Agricultural University Papers 90 (1990) 1–167.

[8] A. Enayati, J. Lines, R. Maharaj, J. Hemingway, Suppressing the vector. In: Shrinking the Malaria Map: A Prospectus on Malaria Elimination, Chapter 9., San Francisco: The Global Health Group, Global Health Sciences, University of California, San Francisco, 2009.

[9] B. Greenwood, Control to elimination: implications for malaria research, Trends in Parasitology 24 (2008) 449–454.

[10] K. Mendis, A. Rietveld, M. Warsame, A. Bosman, B. Greenwood, W. Wernsdorfer, From malaria control to eradication: The WHO perspective, Tropical Medicine and International Health 4 (2009) 802–809.

[11] C. Lengeler, Insecticide-treated bed nets and curtains for preventing malaria., Cochrane Database System Review (2004).

[12] S. Howard, J. Omumbo, E. Some, C. Donnelly, R. Snow, Evidence for a mass community effect of insecticide treated bed nets on the incidence of malaria on the Kenyan Coast., Transactions of The Royal Society of Tropical Medicine and Hygiene 94 (2000) 357–60.

[13] D. Filmer, L. Pritchett, Estimating wealth effects without expenditure data or tears: an application to educational enrollments in states of India., Demography 38 (2001) 115–32.

[14] W. Hawley, F. Philips-Howard, PA ter Kuile, D. Terlouw, J. Vulule, M. Ombok, B. Nahlen, J. Gimnig, S. Kariuki, M. Kolczak, A. Hightower, Community-wide effects of permethrin-treated bed nets on child mortality and malaria morbidity in Western Kenya., American Journal of Tropical Medicine and Hygiene 68 (2003) 121–27.

[15] C. Lengeler, J. Armstrong-Schellenberg, U. D’Alessandro, F. Binka, J. Cattani, Relative versus absolute risk of dying reduction after using insecticide-treated nets for malaria control in Africa., Tropical Medicine & International Health 3 (1998) 286–90.

[16] S. Bhatt, D. Weiss, E. Cameron, D. Bisanzio, B. Mappin, U. Dalrymple, K. Battle, C. Moyes, A. Henry, P. Eckhoff, E. Wenger, B. O, M. Penny, T. Smith, A. Bennett, J. Yukich, T. Eisele, J. Griffin, C. Fergus, M. Lynch, F. Lindgren, J. Cohen, C. Murray, D. Smith, S. Hay, R. Cibulskis, P. Gething, The effect of malaria control on Plasmodium falciparum in Africa between 2000 and 2015., Nature 526 (2015) 207–11.

[17] S. E. Clarke, C. Bøgh, R. C. Brown, M. Pinder, G. E. Walraven, S. W. Lindsay, Do untreated bednets protect against malaria?, Transactions of the Royal Society of Tropical Medicine and Hygiene 95 (2001) 457–462.

[18] G. W. Fegan, M. N. Abdisalan, S. A. Willis, C. Simon, R. Snow, Effect of expanded insecticide-treated bednet coverage on child survival in rural Kenya: a longitudinal study, Lancet 370 (2007) 1035–39.

[19] K. Ernst, M. Hayden, H. Olsen, J. Cavanaugh, I. Ruberto, M. Agawo, S. Munga, Comparing ownership and use of bed nets at two sites with differential malaria transmission in Western Kenya., Malaria Journal 15 (2016) 217.

[20] J. Omumbo, R. Snow, Plasmodium falciparum prevalence in East Africa: a review of empirical data 1927-2003., East African Medical Journal 81 (2004) 649–56.

[21] N. De La Cruz, B. Crookston, K. Dearden, B. Gray, N. Ivins, A. Alder, R. Davis, Who sleeps under bednets in Ghana? a doer/non-doer analysis of malaria prevention behaviours., Malaria Journal 5 (2006) 61.

[22] M. Kayedi, J. Lines, A. Haghdoost, M. Vatandoost, Y. Rassi, K. Khamisabady, Evaluation of the effects of repeated hand washing, sunlight, smoke and dirt on the persistence of deltamethrin on insecticide-treated nets, Transactions of the Royal Society of Tropical Medicine and Hygiene 102 (2008) 811–816.

[23] H. Atieli, D. Menya, A. Githeko, T. Scott, House design modifications reduce indoor resting malaria vector densities in rice irrigation scheme area in Western Kenya, Malaria Journal 8 (2009) 108.

[24] WHO, World Malaria Report 2013 Geneva, WHO Genva, 2013.

[25] W. Brogdon, J. McAllister, Simplification of adult mosquito bioassays through use of time-mortality determinations in glass bottles., Journal of the American Mosquito Control Association 14 (1998.) 159–164.

[26] H. Mawejje, C. Wilding, E. Rippon, A. Hughes, D. Weetman, M. Donnelly, Insecticide resistance monitoring of eld-collected Anopheles gambiae s.l. populations from Jinja, Eastern Uganda, identifies high levels of pyrethroid resistance., Medical and Veterinary Entomology 27 (2013) 276–283.

[27] E. Ochomo, N. Bayoh, E. Walker, B. Abongo, M. Ombok, C. Ouma, A. K. Githeko, J. Vulule, G. Yan, J. E. Gimnig, The efficacy of long-lasting nets with declining physical integrity may be compromised in areas with high levels of pyrethroid resistance., Malaria Journal 12 (2013).

[28] K. H. Toe, C. M. Jones, S. N’Fale, H. M. Ismail, R. K. Dabire, H. Ranson, Increased pyrethroid resistance in malaria vectors and decreased bed net effectiveness, Burkina Faso, Emerging Infectious Diseases 20 (2014) 1691–6.

[29] P. Ojuka, Y. Boum, L. Denoeud-Ndam, C. Nabasumba, Y. Muller, M. Okia, J. D. P. Mwanga-Amumpaire, N. Protopopoff, J. Etard, Early biting and insecticide resistance in the malaria vector Anopheles might compromise the effectiveness of vector control intervention in Southwestern Uganda, Malaria Journal 14 (2015) 148.

[30] N. Moiroux, M. Gomez, C. Pennetier, E. Elanga, Djènontin, F. Chandre, I. Djègbé, G. H, V. Corbel, Changes in Anopheles funestus biting behavior following universal coverage of long-lasting insecticidal nets in Benin, The Journal of Infectious Diseases 206 (2012) 1622–9.

[31] E. committee on insecticides, 1957, URL: http://www.who.int/iris/handle/10665/40380.

[32] G. MacDonald, The epidemiology and control of malaria, London: Oxford University Press, 1957.

[33] R. Fontaine, Integrated mosquito control methodologies; The use of residual insecticides for the control of adult mosquitoes, pp. 49–81, volume 1, London: Academic Press, 1983.

[34] N. Gratz, The future of vector biology and control in the World Health Organization, Journal of the American Mosquito Control Association 1 (1985) 273–278.

[35] J. Hemingway, L. Field, J. Vontas, An overview of insecticide resistance., Science 298 (2002) 96–97.

[36] F. Plapp, Biochemical genetics of insecticide resistance, Annual Review of Entomology 21 (1976) 179–197.

[37] T. Awolola, O. Oduola, C. Strode, L. Koekemoer, B. Brooke, H. Ranson, Evidence of multiple pyrethroid resistance mechanisms in the malaria vector Anopheles gambiae sensu stricto from Nigeria., Transactions of The Royal Society of Tropical Medicine and Hygiene 103 (2009) 1139–45.

[38] T. Davies, L. Field, P. Usherwood, M. Williamson, DDT, pyrethrins, pyrethroids and insect sodium channels, IUBMB Life 59 (2007) 151–162.

[39] F. Ridl, C. Bass, M. Torrez, D. Govender, V. Ramdeen, L. Yellot, A. Edu, C. Schwabe, P. Mohloai, R. Maharaj, I. Kleinschmidt, A pre-intervention study of malaria vector abundance in Rio Muni, Equatorial Guinea: Their role in malaria transmission and the incidence of insecticide resistance alleles., Malaria Journal 7 (2008) 194.

[40] B. Sharp, F. Ridl, D. Govender, J. Kuklinski, I. Kleinschmidt, Malaria vector control by indoor residual insecticide spraying on the tropical island of Bioko, Equatorial Guinea., Malaria Journal 6 (2007) 52.

[41] J. Vontas, G. Small, J. Hemingway, Glutathione S-transferases as antioxidant defence agents confer pyrethroid resistance in Nilaparvata lugens., Biochemical Journal 357 (2001) 65–72.

[42] C. Bogh, E. Pedersen, D. Mukoko, J. Ouma, Permethrin-impregnated bednet effects on resting and feeding behaviour of lymphatic filariasis vector mosquitoes in kenya., Medical and Veterinary Entomology 12 (1998) 52–59.

[43] C. Garrett-Jones, C. Pant, Feeding habits of Anophelines (Diptera: Culicidae) in 1971-1978, with reference to the human blood index: a review., Bulletin of entomological research 70 (1980) 165–185.

[44] V. Balabanidou, L. Grigoraki, J. Vontas, Insect cuticle: a critical determinant of insecticide resistance, Current opinion in insect science 27 (2018) 68–74.

[45] C. Mulamba, J. Riveron, S. Ibrahim, H. Irving, K. Barnes, L. Mukwaya, J. Birungi, C. S. Wondji, Widespread pyrethroid and ddt resistance in the major malaria vector Anopheles funestus in East Africa is driven by metabolic resistance mechanisms., PLoS One 9 (2014) 110058.

[46] C. Edi, B. Koudou, C. Jones, D. Weetman, H. Ranson, Multiple-insecticide resistance in Anopheles gambiae mosquitoes, Southern Cote d’Ivoire, Emerg Infect Dis. 18 (2012) 1508–11.

[47] J. Utzinger, Y. Tozan, B. Singer, Efficacy and cost-effectiveness of environmental management for malaria control, Tropical Medicine and International Health 6 (2001) 677–687.

[48] B. Brooke, Can a single mutation produce an entire insecticide resistance phenotype?, Transactions of The Royal Society of Tropical Medicine and Hygiene 102 (2008) 524–525.

[49] R. Ross, The prevention of malaria, J. Murray, 1910.

[50] G. Macdonald, et al., The analysis of infection rates in diseases in which super infection occurs., Tropical Diseases Bulletin 47 (1950) 907–915.

[51] K. Dietz, L. Molineaux, A. Thomas, A malaria model tested in the african savannah, Bulletin of the World Health Organization 50 (1974) 347.

[52] G. A. Ngwa, W. S. Shu, A mathematical model for endemic malaria with variable human and mosquito populations, Mathematical and Computer Modelling 32 (2000) 747–764.

[53] N. Chitnis, J. M. Cushing, J. Hyman, Bifurcation analysis of a mathematical model for malaria transmission, SIAM Journal on Applied Mathematics 67 (2006) 24–45.

[54] N. Chitnis, J. M. Hyman, J. M. Cushing, Determining important parameters in the spread of malaria through the sensitivity analysis of a mathematical model, Bulletin of Mathematical Biology 70 (2008) 1272.

[55] M. I. Teboh-Ewungkem, C. N. Podder, A. B. Gumel, Mathematical study of the role of gametocytes and an imperfect vaccine on malaria transmission dynamics, Bulletin of Mathematical Biology 72 (2010) 63–93.

[56] J. Arino, A. Ducrot, P. Zongo, A metapopulation model for malaria with transmission-blocking partial immunity in hosts, Journal of Mathematical Biology 64 (2012) 423–448.

[57] M. I. Teboh-Ewungkem, G. A. Ngwa, C. N. Ngonghala, Models and proposals for malaria: a review, Mathematical Population Studies 20 (2013) 57–81.

[58] C. N. Ngonghala, G. A. Ngwa, M. I. Teboh-Ewungkem, Periodic oscillations and backward bifurcation in a model for the dynamics of malaria transmission, Mathematical Biosciences 240 (2012) 45–62.

[59] C. N. Ngonghala, M. I. Teboh-Ewungkem, G. A. Ngwa, Persistent oscillations and backward bifurcation in a malaria model with varying human and mosquito populations: implications for control, Journal of Mathematical Biology 70 (2015) 1581–1622.

[60] C. N. Ngonghala, J. Mohhamed-Awel, R. C. Zhao, O. Porsper, Interplay between insecticide-treated bed-nets and mosquito demography: implications for malaria control, Journal of Theoretical Biology 97 (2016) 179–192.

[61] C. N. Ngonghala, M. I. Teboh-Ewungkem, G. A. Ngwa, Observance of period-doubling bifurcation and chaos in an autonomous ODE model for malaria with vector demography, Theoretical Ecology 9 (2016) 337–351.

[62] S. E. Eikenberry, A. B. Gumel, Mathematical modeling of climate change and malaria transmission dynamics: a historical review, Journal of Mathematical Biology 77 (2018) 857–933.

[63] K. Okuneye, S. E. Eikenberry, A. B. Gumel, Weather-driven malaria transmission model with gonotrophic and sporogonic cycles, Journal of biological dynamics 13 (2019) 288–324.

[64] G. F. Killeen, T. A. Smith, H. M. Ferguson, H. Mshinda, S. Abdulla, C. Lengeler, S. P. Kachur, Preventing childhood malaria in africa by protecting adults from mosquitoes with insecticide-treated nets, PLoS Medicine 4 (2007).

[65] N. Chitnis, A. Schapira, T. Smith, R. Steketee, Comparing the effectiveness of malaria vector-control interventions through a mathematical model, The American Journal of Tropical Medicine and Hygiene 83 (2010) 230–240.

[66] L. C. Okell, L. S. Paintain, J. Webster, K. Hanson, J. Lines, From intervention to impact: modelling the potential mortality impact achievable by different long-lasting, insecticide-treated net delivery strategies, Malaria Journal 11 (2012) 327.

[67] O. J. Briët, D. Hardy, T. A. Smith, Importance of factors determining the effective lifetime of a mass, long-lasting, insecticidal net distribution: a sensitivity analysis, Malaria Journal 11 (2012) 20.

[68] F. Agusto, S. Valle, K. Blayneh, C. Ngonghala, M. Goncalves, N. Li, R. Zhao, H. Gong, The impact of bed-net use on malaria prevalence., Journal of Theoretical Biology 320 (2013) 58–65.

[69] C. N. Ngonghala, S. Y. Del Valle, R. Zhao, J. Mohammed-Awel, Quantifying the impact of decay in bed-net efficacy on malaria transmission, Journal of Theoretical Biology 363 (2014) 247–261.

[70] S. Barbosa, I. M. Hastings, The importance of modelling the spread of insecticide resistance in a heterogeneous environment: the example of adding synergists to bed nets, Malaria Journal 11 (2012) 258.

[71] P. L. Birget, J. C. Koella, A genetic model of the effects of insecticide-treated bed nets on the evolution of insecticide-resistance, Evolution, Medicine, and Public Health 2015 (2015) 205–215.

[72] J. Wairimu, F. Chirove, M. Ronoh, D. M. Malonza, Modeling the effects of insecticides resistance on malaria vector control in endemic regions of Kenya, Biosystems 174 (2018) 49–59.

[73] S. Barbosa, K. Kay, N. Chitnis, I. M. Hastings, Modelling the impact of insecticide-based control interventions on the evolution of insecticide resistance and disease transmission, Parasites & Vectors 11 (2018) 482.

[74] J. Mohammed-Awel, F. Agusto, R. E. Mickens, A. B. Gumel, Mathematical assessment of the role of vec-tor insecticide resistance and feeding/resting behavior on malaria transmission dynamics: Optimal control analysis, Infectious Disease Modelling 3 (2018) 301–321.

[75] F. Wat’senga, E. Z. Manzambi, A. Lunkula, R. Mulumbu, T. Mampangulu, N. Lobo, A. Hendershot, C. Fornadel, D. Jacob, M. Niang, et al., Nationwide insecticide resistance status and biting behaviour of malaria vector species in the Democratic Republic of Congo, Malaria Journal 17 (2018) 129.

[76] G. Fuseini, R. N. Nguema, W. P. Phiri, O. T. Donfack, C. Cortes, M. E. Von Fricken, J. I. Meyers, I. Kleinschmidt, G. A. Garcia, C. Maas, et al., Increased biting rate of insecticide-resistant Culex mosquitoes and community adherence to IRS for malaria control in urban Malabo, Bioko Island, Equatorial Guinea, Journal of Medical Entomology 56 (2019) 1071–1077.

[77] World Bank Data, Life expectancy at birth, total (years) - Sub-Saharan Africa (Accessed on January 3, 2020).

[78] T. world fact book, Central Intelligence Agency. (accessed 27.03.14) Life expectancy at birth, Central Intelligence Agency, 2014.

[79] N. Chitnis, J. M. Hyman, J. M. Cushing, Determining important parameters in the spread of malaria through the sensitivity analysis of a mathematical model., Bulletin of Mathematical Biology 70 (2008) 1272–1296.

[80] Srinivas, immunity, 2015. URL: http://www.malariasite.com/immunity.

[81] M. Teboh-Ewungkem, Malaria control: the role of local communities as seen through a mathematical model in a changing population-cameroon, 2009.

[82] M. Teboh-Ewungkem, J. Tchuenche, Z. Mukandavire, Malaria control: the role of local communities as seen through a mathematical model in a changing population-Cameroon., Advances in Disease Epidemiology. Nova Science Publishers; New York (2009) 101–138.

[83] W. Wang, X.-Q. Zhao, Threshold dynamics for compartmental epidemic models in periodic environments, Journal of Dynamics and Differential Equations 20 (2008) 699–717.

[84] G. Davidson, C. Draper, Field studies of some of the basic factors concerned in the transmission of malaria, Transactions of the Royal Society of Tropical Medicine and Hygiene 47 (1953) 522–535.

[85] H. Giles, D. Warrel, Bruce chwatts’ essential of malariology. 3”‘edition, Edward Arnold, London (1993) 124–163.

[86] J. Lines, J. Myamba, C. Curtis, Experimental hut trials of permethrin-impregnated mosquito nets and eave curtains against malaria vectors in tanzania, Medical and veterinary entomology 1 (1987) 37–51.

[87] M. F. Boyd, Epidemiology: factors related to the definitive host, Malariology 1 (1949) 608–697.

[88] M. Smalley, R. Sinden, Plasmodium falciparum gametocytes: their longevity and infectivity, Parasitology 74 (1977) 1–8.

[89] J. Nedelman, Inoculation and recovery rates in the malaria model of dietz, molineaux, and thomas, Mathematical Biosciences 69 (1984) 209–233.

[90] J. Nedelman, Introductory review some new thoughts about some old malaria models, Mathematical Biosciences 73 (1985) 159–182.

[91] L. Molineaux, G. Shidrawi, J. Clarke, J. Boulzaguet, T. Ashkar, Assessment of insecticidal impact on the malaria mosquito’s vectorial capacity, from data on the man-biting rate and age-composition, Bulletin of the World Health Organization 57 (1979) 265.

[92] S. Gupta, J. Swinton, R. M. Anderson, Theoretical studies of the effects of heterogeneity in the parasite population on the transmission dynamics of malaria, Proceedings of the Royal Society of London. Series B: Biological Sciences 256 (1994) 231–238.

[93] J. Mwangangi, E. Muturi, S. Muriu, J. Nzovu, J. Midega, C. Mbogo, The role of Anopheles arabiensis and Anopheles coustani in indoor and outdoor malaria transmission in Taveta District, Kenya, Parasites and Vectors 6 (2013) 114.

[94] P. Van den Driessche, J. Watmough, Reproduction numbers and sub-threshold endemic equilibria for compartmental models of disease transmission, Mathematical Biosciences 180 (2002) 29–48.

[95] B. Ondeto, C. Nyundo, L. Kamau, S. Muriu, J. M. Mwangangi, K. Njagi, E. M. Mathenge, H. Ochanda, C. Mbogo, Current status of insecticide resistance among malaria vectors in Kenya, Parasites & Vectors 10 (2017) 429.

